# Rare genetic variation in VE-PTP is associated with central serous chorioretinopathy, venous dysfunction and glaucoma

**DOI:** 10.1101/2024.05.08.24307013

**Authors:** Joel T Rämö, Bryan Gorman, Lu-Chen Weng, Sean J Jurgens, Panisa Singhanetr, Marisa G Tieger, Elon HC van Dijk, Christopher W Halladay, Xin Wang, Joost Brinks, Seung Hoan Choi, Yuyang Luo, FinnGen, Program VA Million Veteran, Saiju Pyarajan, Cari L Nealon, Michael B Gorin, Wen-Chih Wu, Lucia Sobrin, Kai Kaarniranta, Suzanne Yzer, Aarno Palotie, Neal S Peachey, Joni A Turunen, Camiel JF Boon, Patrick T Ellinor, Sudha K Iyengar, Mark J Daly, Elizabeth J Rossin

## Abstract

Central serous chorioretinopathy (CSC) is a fluid maculopathy whose etiology is not well understood. Abnormal choroidal veins in CSC patients have been shown to have similarities with varicose veins. To identify potential mechanisms, we analyzed genotype data from 1,477 CSC patients and 455,449 controls in FinnGen. We identified an association for a low-frequency (AF=0.5%) missense variant (rs113791087) in the gene encoding vascular endothelial protein tyrosine phosphatase (VE-PTP) (OR=2.85, P=4.5×10^-9^). This was confirmed in a meta-analysis of 2,452 CSC patients and 865,767 controls from 4 studies (OR=3.06, P=7.4×10^-15^). Rs113791087 was associated with a 56% higher prevalence of retinal abnormalities (35.3% vs 22.6%, P=8.0x10^-4^) in 708 UK Biobank participants and, surprisingly, with varicose veins (OR=1.31, P=2.3x10^-11^) and glaucoma (OR=0.82, P=6.9x10^-9^). Predicted loss-of-function variants in VEPTP, though rare in number, were associated with CSC in All of Us (OR=17.10, P=0.018). These findings highlight the significance of VE-PTP in diverse ocular and systemic vascular diseases.

## Introduction

Central serous chorioretinopathy (CSC) is a maculopathy associated with a thickened and dilated choroidal vasculature, retinal pigment epithelium (RPE) detachments and subretinal fluid (SRF). CSC often first manifests with decreased visual acuity in individuals between 30 and 50 years of age^1^. CSC is relatively common worldwide, with annual age-adjusted incidence estimates between 5.8:100,000 and 34:100,000 in different populations^1,2^; a prevalence as high as 1.7% has been reported in India^3,4^.

For some patients the SRF resolves spontaneously and quickly, but for others the SRF persists or recurs leading to photoreceptor damage and visual decline. Photodynamic therapy is a preferred treatment for chronic CSC but is ineffective in resolving SRF in a noteworthy percentage of patients^2^.

The etiology of CSC is not well understood, limiting therapeutic development and primary prevention. Imaging of the choroidal vessels via fluorescein and indocyanine green (ICG) angiography have revealed characteristic focal leakage and staining patterns representing leakage of dye from the choroid to the subretinal space^5,6^. Several risk factors have been noted, including endogenous and exogenous corticosteroids and pregnancy^7^; however, none of these have a clear known pathophysiologic mechanism causing CSC.

Original etiologic discussions focused on RPE hyperpermeability, but this theory does not explain the notably thickened choroid with dilated vasculature. Spaide and colleagues brought forth a convincing mechanistic explanation in 2022 which incorporated recent enhanced depth optical coherence tomography (OCT) and widefield ICG angiography findings and ascribed CSC at least in part to intrinsic venous outflow insufficiency^8^. The dilated vasculature, delayed choroidal filling and arterio-venous anastomoses noted on ICG angiography contribute toward a theory of CSC being a venous overload choroidopathy. The vascular remodeling in CSC has been compared most closely to varicose veins, although the changes are on a much smaller scale. Varicose veins are a common disease affecting approximately 20% of the population (females more than males) and represent a weakening of the vessel wall with resultant vascular dilation in the lower extremity (typically in the greater and lesser saphenous veins)^9^.

Genome-wide analysis of variants associated with CSC has recently enjoyed success^10–12^. To date, these studies have identified 6 common variant loci linked to CSC. In a recent CSC meta-analysis^12^, we identified associated loci encoding genes that are preferentially expressed in the choroidal vasculature, which complements the theory of CSC being a disorder of vascular competence. However, aside from the well-documented Complement Factor H locus—shared between CSC and age-related macular degeneration (AMD)^10–13^—no coding variants or clearly causal non-coding variants have robustly pointed to specific genes, and thus mechanistic understanding remains limited.

Here, we conduct a genome-wide association study of CSC in new data from the FinnGen study. We further evaluate the associations of a newly identified locus with CSC and co-associated traits across 5 studies including over 1.3 million individuals. These findings provide an intriguing window into shared underlying pathophysiology between ocular diseases and systemic vascular dysfunction.

## Results

### A genome-wide association study of CSC in FinnGen identifies a missense variant in *PTPRB*

We began by conducting a case-control GWAS including 1,477 patients with CSC and 455,449 controls in FinnGen (**Supplementary Table 1**; **Supplementary Results**). The prevalence of other ocular diseases among patients with CSC was low (**Supplementary Table 2 and Supplementary Results**). This analysis identified 3 loci at genome-wide significance (**Figure 1** and **Supplementary Figure 2**), 2 of which have been previously reported and are marked by common noncoding variants at *CFH* (Complement Factor H) and *CD46* (Membrane cofactor protein). At the 12q15 locus, the lead variant was a low-frequency (AF = 0.5%) missense variant (rs113791087, 12:70559589:T:G) in the *PTPRB* (Protein Tyrosine Phosphatase Receptor Type B; NCBI Gene ID: 5787) gene that encodes the VE-PTP protein (UniProt ID: P23467) (**Supplementary Figure 3**). Rs113791087 was associated with increased risk of CSC rs113791087 (OR = 2.85 [2.01-4.05] per G allele, P = 4.5×10^-9^). Fine-mapping with the sum of single effects (SuSiE) approach nominated rs113791087 as the most likely causal variant in the locus, with a posterior inclusion probability of 0.995 in its credible set.

**Figure 1.**
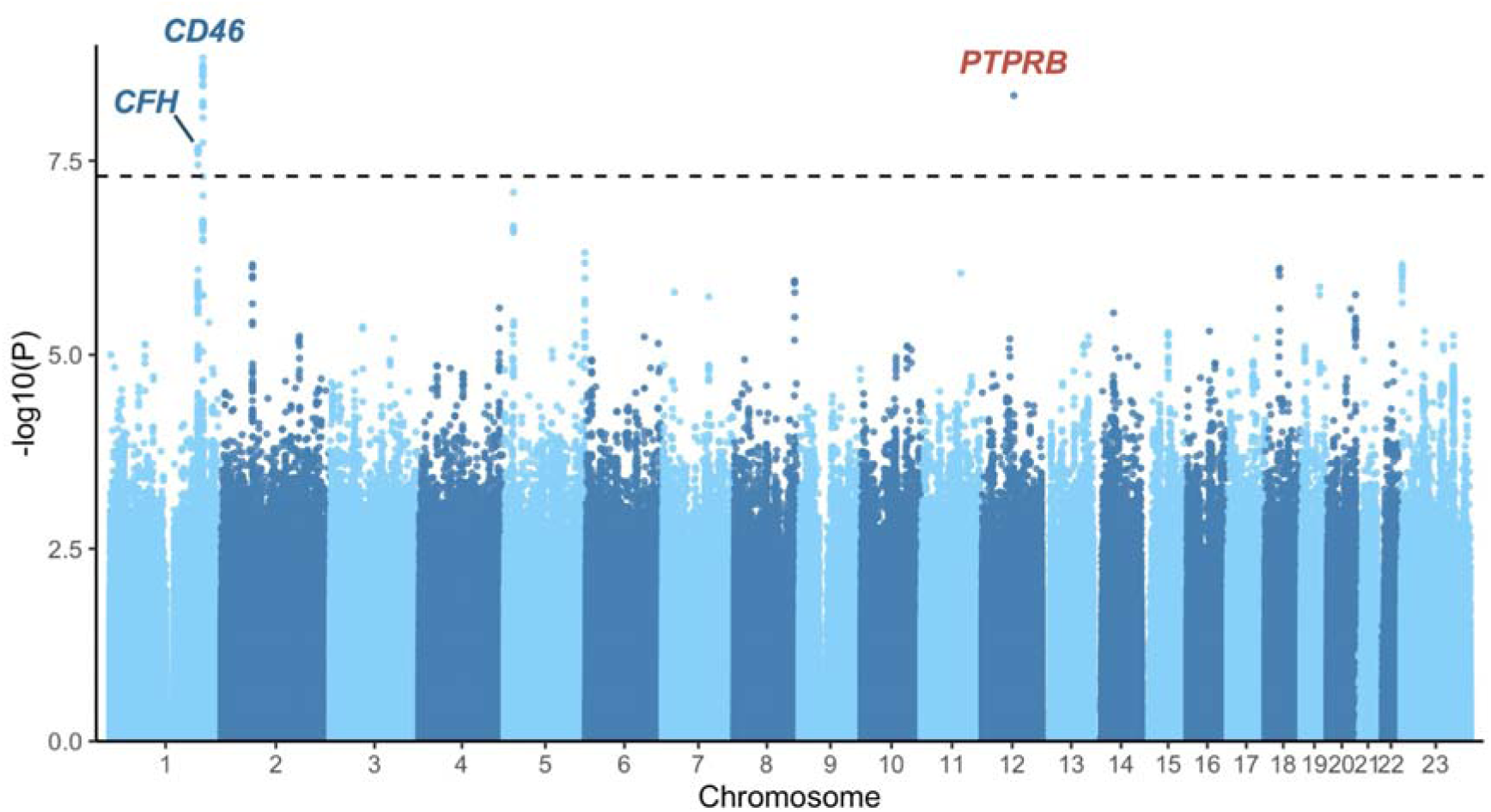
Genome-wide association study of central serous chorioretinopathy in FinnGen. A genome-wide association study of central serous chorioretinopathy was conducted including 1,477 patients with central serous chorioretinopathy and 455,449 controls from the FinnGen study. Each genomic variant is plotted as a data point, with P-values shown on the y-axis on a logarithmic scale and chromosomal position shown on the x-axis. The genome-wide significance threshold (P = 5×10^-8^) is shown with a dashed line. In each of three loci reaching genome-wide significance, the nearest protein-coding gene to the lead variant is labeled (blue = previously reported loci, red = novel locus).

To ensure that this association was not confounded by AMD—which has certain overlapping phenotypic and genotypic features with CSC—we conducted a sensitivity analysis by excluding patients with AMD symmetrically from both CSC patients and controls and observed that the association of rs113791087 with CSC remained consistent (1092 patients and 485392 controls, OR = 2.95 [1.97-4.43], P = 1.8×10^-7^) (**Supplementary Table 4**). The effect size remained robust even after excluding 37 diagnosis codes that reflect potentially confounding causes of fluid maculopathy from patients and controls (769 patients and 452038 controls, OR = 3.15 [1.97-5.05], P = 1.8×10^-6^) (**Supplementary Tables 3-4**)^14^.

### Replication and meta-analysis of the association of *PTPRB* rs113791087 with CSC

We carried out replication of variants in the *PTPRB* locus in 3 independent studies, including newly analyzed data from Million Veteran Program (MVP) participants of European ancestry (706 patients with CSC and 273,198 controls, **Supplementary Table 1**), newly analyzed data from All of Us participants of European ancestry (133 patients with CSC and 103,600 controls, **Supplementary Table 1**), and data from a previously reported European chronic CSC cohort (521 patients with chronic CSC and 3,577 population controls)^10^. Patients with AMD were excluded from all newly analyzed studies contributing to the meta-analysis and had been previously excluded from analyses of the chronic CSC cohort^10^. We observed consistently significant associations between rs113791087 and CSC in all studies (**Figure 2** and **Supplementary Table 4**) and in a cross-study meta-analysis of the locus (OR = 3.06 [95% CI 2.31-4.06], P = 7.3×10^-15^; 2452 patients with CSC and 865767 controls) (**Supplementary Figure 4**). Additionally, rs113791087 was the lead variant in its locus in the All of Us study even when considering all variants with a minor allele count > 40 uncovered by whole genome sequencing (**Supplementary Figure 5**). The point estimate was higher in All Of Us (OR = 8.28 [95% CI 3.37-20.33]; 133 patients with CSC and 103600 controls) compared with studies that included more patients with CSC, but confidence intervals were overlapping between all studies. All 133 patients with CSC in All of Us were unrelated.

**Figure 2.**
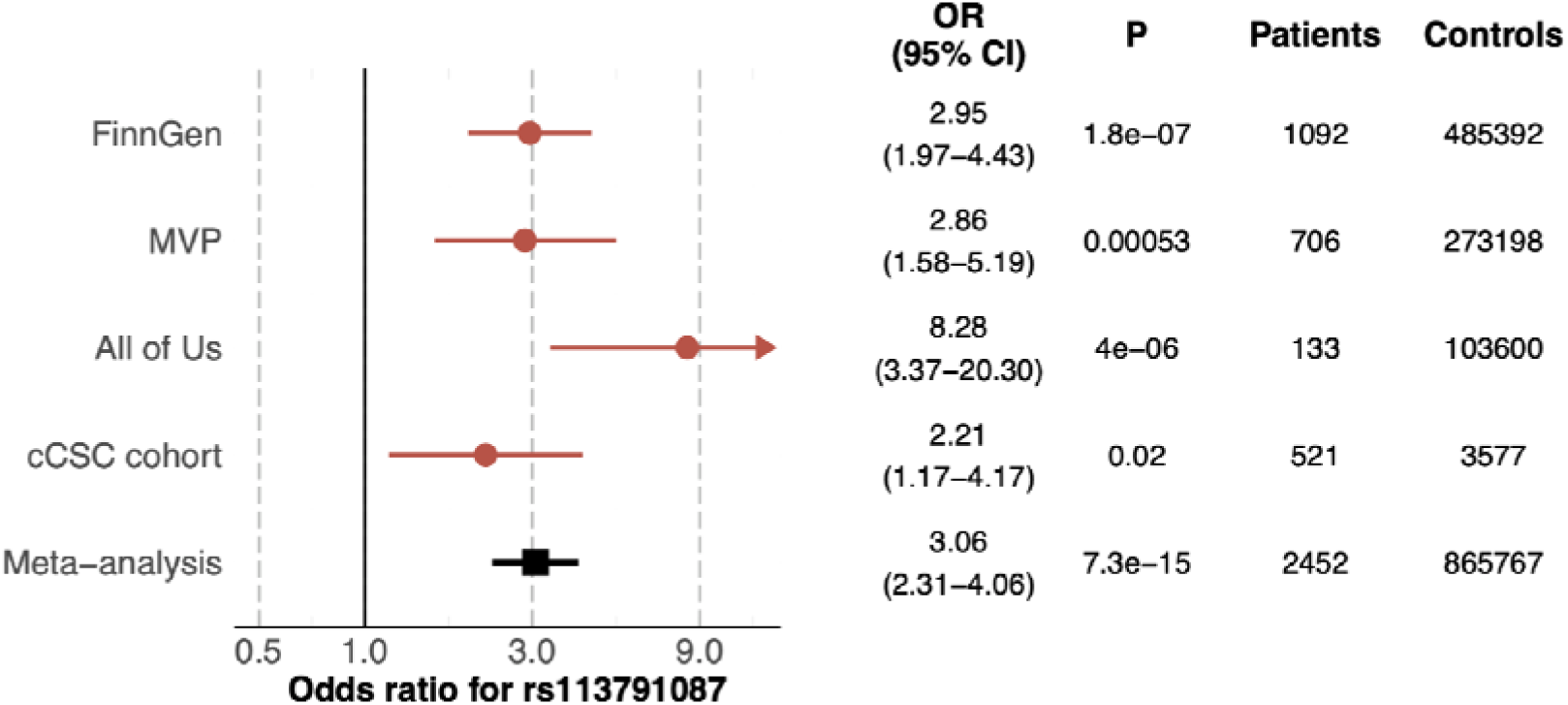
Associations of the *PTPRB* missense variant rs113791087 with central serous chorioretinopathy in 4 studies. The association of rs113791087 with central serous chorioretinopathy (CSC) was examined in 4 different studies. In all biobank-based studies (FinnGen, Million Veteran Program [MVP] and All of Us) included in the meta-analysis, patients with CSC were identified based on International Statistical Classification of Diseases codes, and all participants with age-related macular degeneration were excluded from patients and controls following a harmonized study protocol. In the chronic CSC (cCSC) cohort, patients were identified from ophthalmological clinics based on expert review. For the All of Us cohort, the upper range of the confidence interval is truncated (arrow). An inverse-weighted fixed-effects meta-analysis was conducted to combine data from all studies. We observed no statistically significant heterogeneity for rs113791087 in the meta-analysis (I^2^ = 0.48, Q-statistic = 5.8, Q-statistic P-value = 0.12).

In addition to the broad sensitivity analysis conducted in FinnGen, the granularity of ICD-10-CM and ICD-9-CM codes in MVP allowed us to examine the association of rs113791087 with dystrophies primarily involving the RPE (H35.54 and 362.76, respectively), an endpoint that includes pattern dystrophy which can pose diagnostic challenge with respect to CSC. We observed no significant association of rs113791087 with this outcome (OR = 1.20 [0.78– 1.86], P = 0.41; 2228 patients and 311134 controls).

We also evaluated another missense variant in the *PTPRB* gene (rs61758735, 12:70555234:G>A, AF = 0.69%), which has previously been suggestively associated with CSC in 2 Dutch families in an exome sequencing study and which is in linkage equilibrium (D’ = 1.0, R^2^ = 0.0) with rs113791087 according to 1000 Genomes European subpopulation reference panel^15,16^. In the meta-analysis of 4 studies, rs61758735 was associated with increased risk of CSC at nominal significance although with a smaller effect size than rs113791087 (OR = 1.63 [1.15–2.32], P = 0.006142).

### Rs113791087 is associated with retinal abnormalities on optical coherence tomography

Genetic risk may manifest in subclinical differences even in the absence of diagnosed disease. To identify retinal characteristics potentially associated with the rs113791087 variant, we extracted optical coherence tomography (OCT) images from 266 participants who were heterozygous or homozygous for the G allele of rs113791087 and 442 age-matched participants with the TT allele in UK Biobank (UKB). These images were graded by 3 ophthalmologists blinded to genotype status. We observed an increased overall prevalence of retinal abnormalities at the level of the RPE in participants with the GT or GG genotype compared to participants with the TT genotype (35.3% vs 22.6%, P=0.00078 corrected for age, sex, examiner and first 10 PCs) (**Figure 3** and **Supplementary Table 5**).

**Figure 3.**
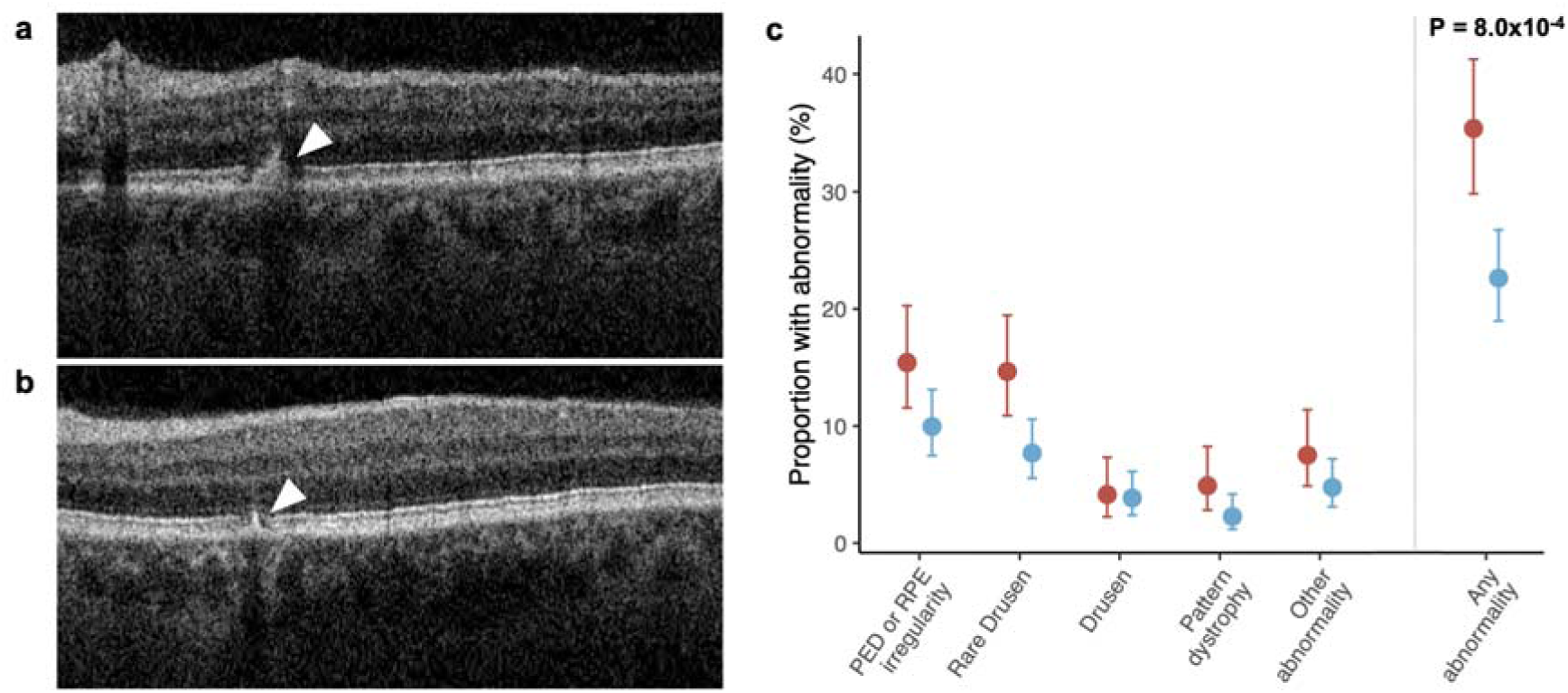
Retinal abnormalities in optical coherence tomography images by *PTPRB* rs113791087 genotype status in UK Biobank. Optical coherence tomography (OCT) images of 266 participants with the rs113791087 G allele (GG or GT genotype) and 442 age-matched participants lacking the G allele (TT genotype) were obtained from UKB. Images were independently evaluated by 3 retina specialists who were blinded to genotype. Each grader was tasked with identifying and categorizing retinal pigment epithelium (RPE) abnormalities according to the following categories: rare drusen (1-5), drusen (>5), pattern dystrophy, pigment epithelial detachment (PED) or nonspecific RPE irregularity, and other rare findings (subretinal fluid, pachychoroid pigment epitheliopathy, atrophy, intraretinal fluid, or evidence of central serous chorioretinopathy). Panel **a** depicts a B-scan superior to the fovea showing an area of RPE irregularity and pigment migration (arrowhead) from a randomly selected participant, who had the TT genotype. Panel **b** depicts a B-scan inferior to the fovea showing a second area of RPE irregularity and pigment migration (arrowhead) in the same participant. Panel **c** shows the prevalence of abnormalities for participants with the GG or GT genotype (red) and TT genotype (blue). Statistical significance was evaluated using linear regression including age, sex, examiner and the first 10 PCs as covariates. Binomial proportion 95% confidence intervals were calculated using the Agresti-Coull method. Optical coherence tomography images are reproduced by kind permission from UK Biobank ©.

### Co-associations of *PTPRB* rs13791087 with varicose veins and glaucoma revealed by a phenome-wide association study in FinnGen

We conducted a phenome-wide association study of 2,469 phenotypes in FinnGen to identify potential pleiotropic effects of the *PTPRB* rs113791087 variant. We observed a genome-wide significant association with varicose veins of the lower extremity (38,467 patients and 432,223 controls, OR = 1.39 [1.25-1.54], P = 3.1x10^-10^) (**Figure 4** and **Supplementary Table 6**). Among the most significant suggestive-level associations, we noted an increased risk of venous thromboembolism (OR = 1.29 [1.15-1.45], P = 1.8x10^-5^), an increased risk of pleural effusion (OR = 1.55 [1.24-1.94], P = 9.5x10^-5^), and a reduced risk of glaucoma (OR = 0.79 [0.69-0.90], P = 5.1×10^-4^).

**Figure 4.**
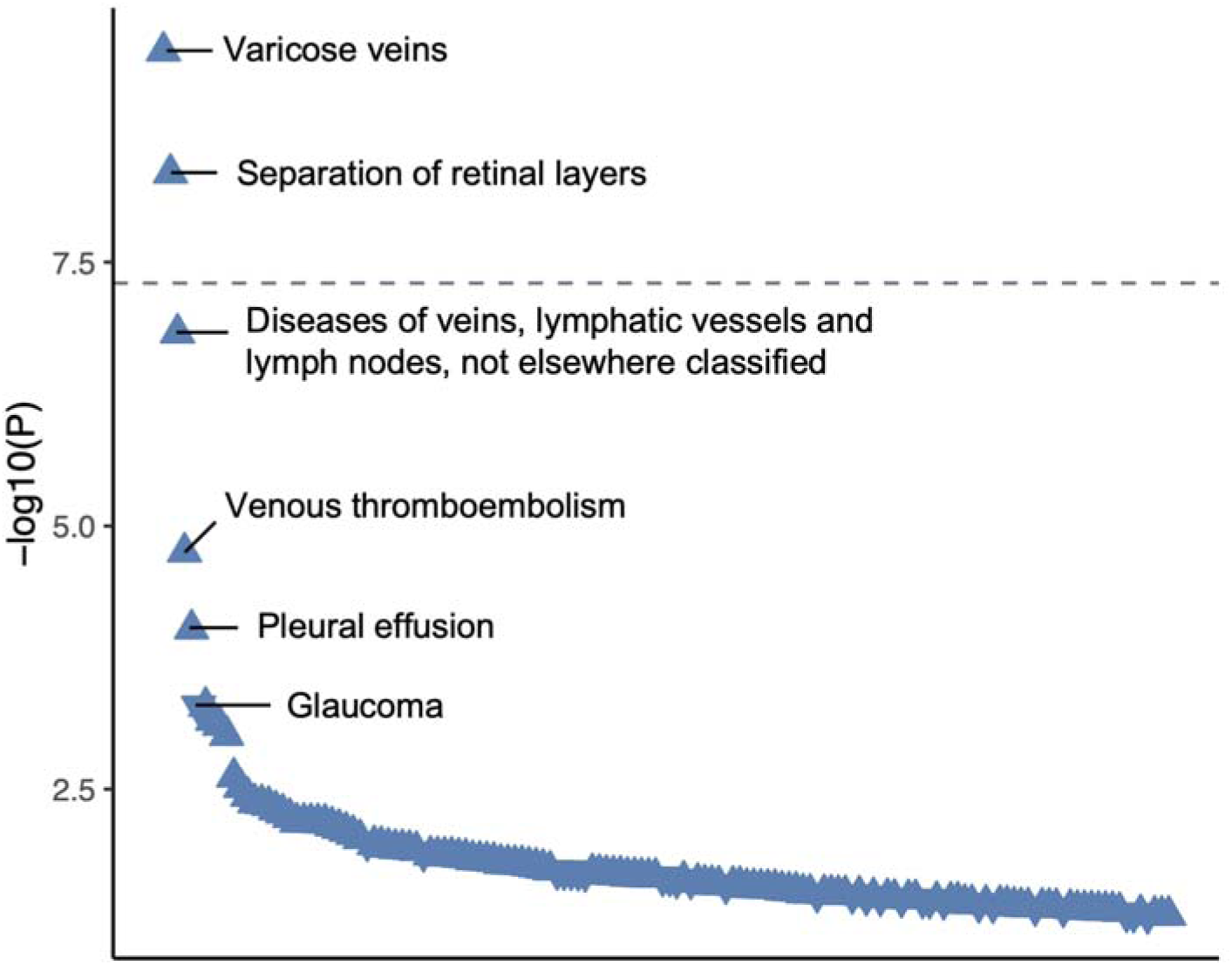
Phenome-wide association study of the *PTPRB* missense variant rs113791087 in FinnGen. To identify potential pleiotropic associations of the rs113791087 variant, a phenome-wide association study was conducted including 2,469 phenotypes. Data are shown for all phenotypes that were at least nominally associated (P < 0.05) with rs113791087. The negative common logarithm of each P-value is shown on the y-axis. The genome-wide significance threshold (P = 5×10^-8^) is shown with a dashed line. The direction of association with the risk of each disease is denoted by symbols (arrow up = increased risk, arrow down = decreased risk).

By contrast, rs113791087 was not significantly associated with common eye diseases including wet AMD (OR = 1.05 [0.81-1.35], P = 0.72), dry AMD (OR = 1.03 [0.82-1.29], P = 0.79), or diabetic retinopathy (OR = 0.98 [0.82-1.18], P = 0.59).

Because VE-PTP has been investigated as a target for diabetic macular edema^17^, we conducted further association analyses limited to participants with type 1 or type 2 diabetes, and observed no significant associations of rs113791087 with diabetic retinopathy among type 1 (OR = 2.14 [95% CI 0.88–5.20], P = 0.092; 2712 patients and 1116 controls) or type 2 diabetic patients (OR = 0.88 [95% CI 0.66–1.16], P = 0.355; 5443 patients and 77435 controls), or with diabetic maculopathy among type 1 (OR = 1.40 [95% CI 0.53–3.71], P = 0.50; 715 patients and 3113 controls) or type 2 diabetic patients (OR = 0.79 [95% CI 0.49– 1.29], P = 0.352; 1724 patients and 81154 controls).

To better understand the co-association of rs113791087 with CSC and varicose veins of the lower extremity, we evaluated the association of these diseases in the general FinnGen population. We observed no difference in the risk of CSC among all participants with varicose veins (OR = 0.99 [95% CI 0.98–1.01], P = 0.43), suggesting that the shared association of rs113791087 with both outcomes may be reflective of true causal associations with both phenotypes rather than simple population-level correlation between the phenotypes.

The clinical manifestations of varicose veins of the lower extremity are broad-ranging and include, among others, asymptomatic venous dilation, edema, and ulceration. In analyses of ICD code based varicose vein subtypes in FinnGen, we observed that rs113791087 was associated with uncomplicated varicose veins of the lower extremity (OR = 1.39 [1.25–1.53], P = 4.3×10^-10^; 38,025 patients and 455,679 controls), but not with ulcerated varicose veins of the lower extremity (OR = 0.93 [0.68–1.28], P = 0.67; 4,075 patients and 455,679 controls).

### Cross-study meta-analyses of most significant disease associations for rs13791087

We evaluated the robustness of associations between rs113791087 and the top 5 distinct diseases from the FinnGen phenome-wide association study by conducting single-variant meta-analyses including data from FinnGen, MVP, UKB, and All of Us (**Figure 5** and **Supplementary Tables 7-11**). In this meta-analysis, rs113791087 was associated with increased risk of uncomplicated varicose veins (OR = 1.31 [95% CI 1.21–1.42], P = 2.3x10^-11^; 62621 patients and 1341326 controls) (**Figure 5** and **Supplementary Table 11)**. We also observed an association between rs113791087 and reduced risk of glaucoma at genome-wide significance (OR = 0.82 [95% CI 0.76–0.88], P = 6.9x10^-9^; 125075 patients and 1290261 controls); the effect estimate was concordant for primary open-angle glaucoma (OR = 0.82 [95% CI 0.72–0.93], P = 0.0014; 35557 patients and 1366253 controls). Consistent with this observation, rs113791087 was also associated with 0.51mmHg [95% CI 0.26–0.76mmHg] lower intraocular pressure (P = 5.2×10^-5^) among 77,449 UKB participants.

**Figure 5.**
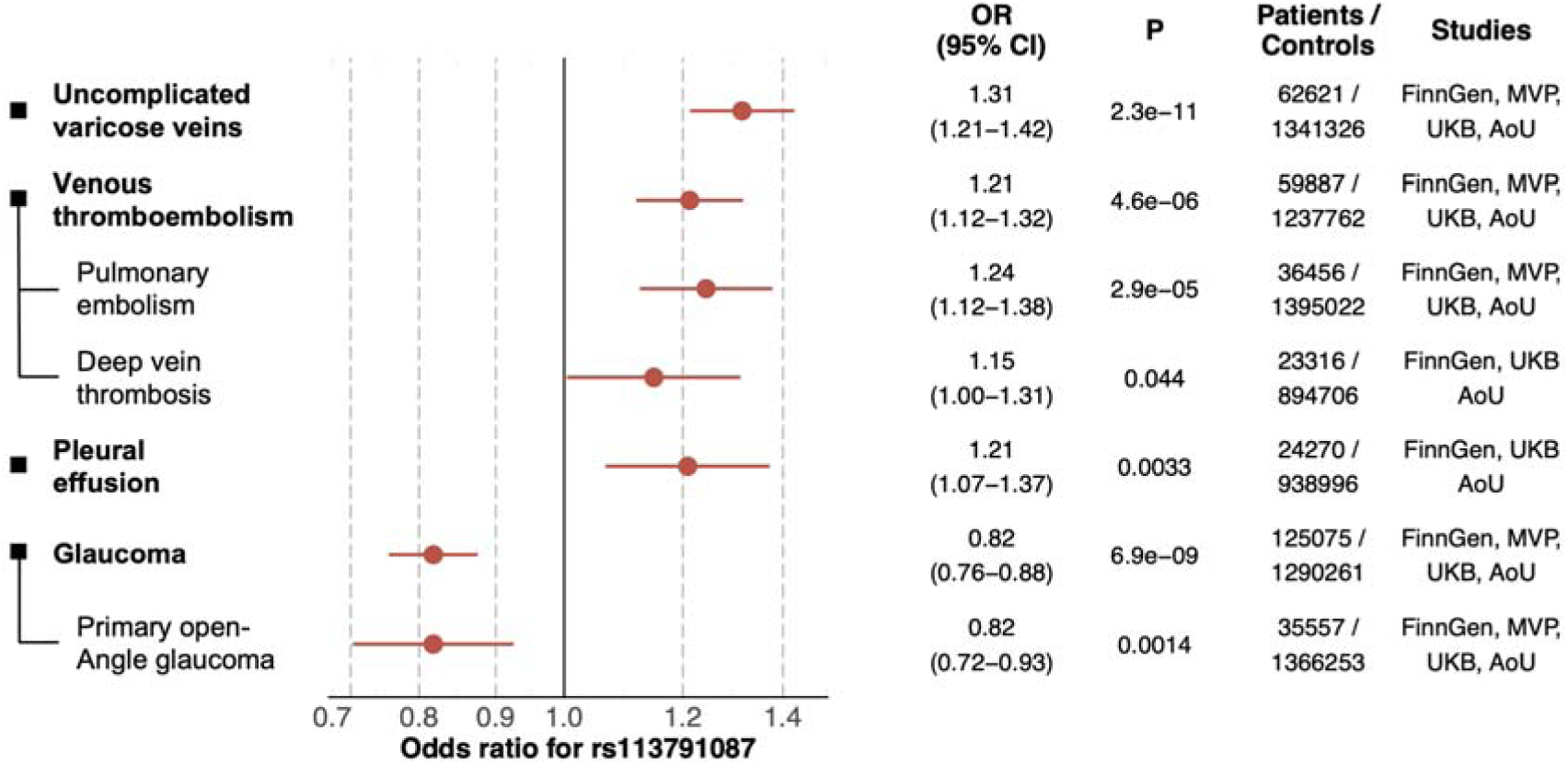
Cross-study meta-analyses of other diseases most significantly associated with the *PTPRB* missense variant rs113791087. The distinct diseases most significantly associated with rs113791087 in the phenome-wide association study of rs113791087 in FinnGen were carried forward for multi-study meta-analyses including available data from FinnGen, UK Biobank (UKB), Million Veteran Program (MVP), and All of Us (AoU). Pulmonary embolism and deep vein thrombosis were examined separately as the major subtypes of venous thromboembolism, and primary open-angle glaucoma was examined separately as a major subtype of glaucoma. Association results from contributing studies were combined in an inverse variance weighted meta-analysis.

In addition to the genome-wide significant associations, in the cross-study meta-analyses we observed a suggestive association between rs113791087 and increased risk of venous thromboembolism (OR = 1.21 [95% 1.12-1.32], P = 4.6x10^-6^; 59887 patients and 1237762 conrols). Association estimates for the constituent endpoints of pulmonary embolism (OR = 1.24 [95% CI 1.12–1.38], P = 2.9x10^-5^; 36456 patients and 1395022 controls) and deep vein thrombosis (OR = 1.15 [95% CI 1.00–1.31], P = 0.044; 23316 patients and 894706 controls) were directionally concordant.

### Disease associations of predicted rare loss-of-function variants in *PTPRB*

To identify other genetic variation in *PTPRB* that might be associated with ocular or vascular systemic diseases, and to inform whether rs113791087 might act in a loss-of-function or gain-of-function manner, we identified participants who had rare pLOF *PTPRB* variants in UKB (n = 120 participants with pLOF variant) and All of Us (n = 75 participants with pLOF variant). While statistical power was limited, among traits highlighted by the rs113791087 phenome-wide association study, participants with a *PTPRB* pLOF variant had suggestively increased risk of CSC (OR = 17.10 [95% CI 1.64–178.00], P = 0.018), venous thromboembolism (OR = 2.22 [95% CI 1.29–3.83], P = 0.0039), pulmonary embolism (OR = 2.90 [95% 1.51–5.57], P = 0.0014), and deep vein thrombosis (OR = 2.15 [95% CI 1.08– 4.27], P = 0.029) (**Figure 6** and **Supplementary Table 12**).

**Figure 6.**
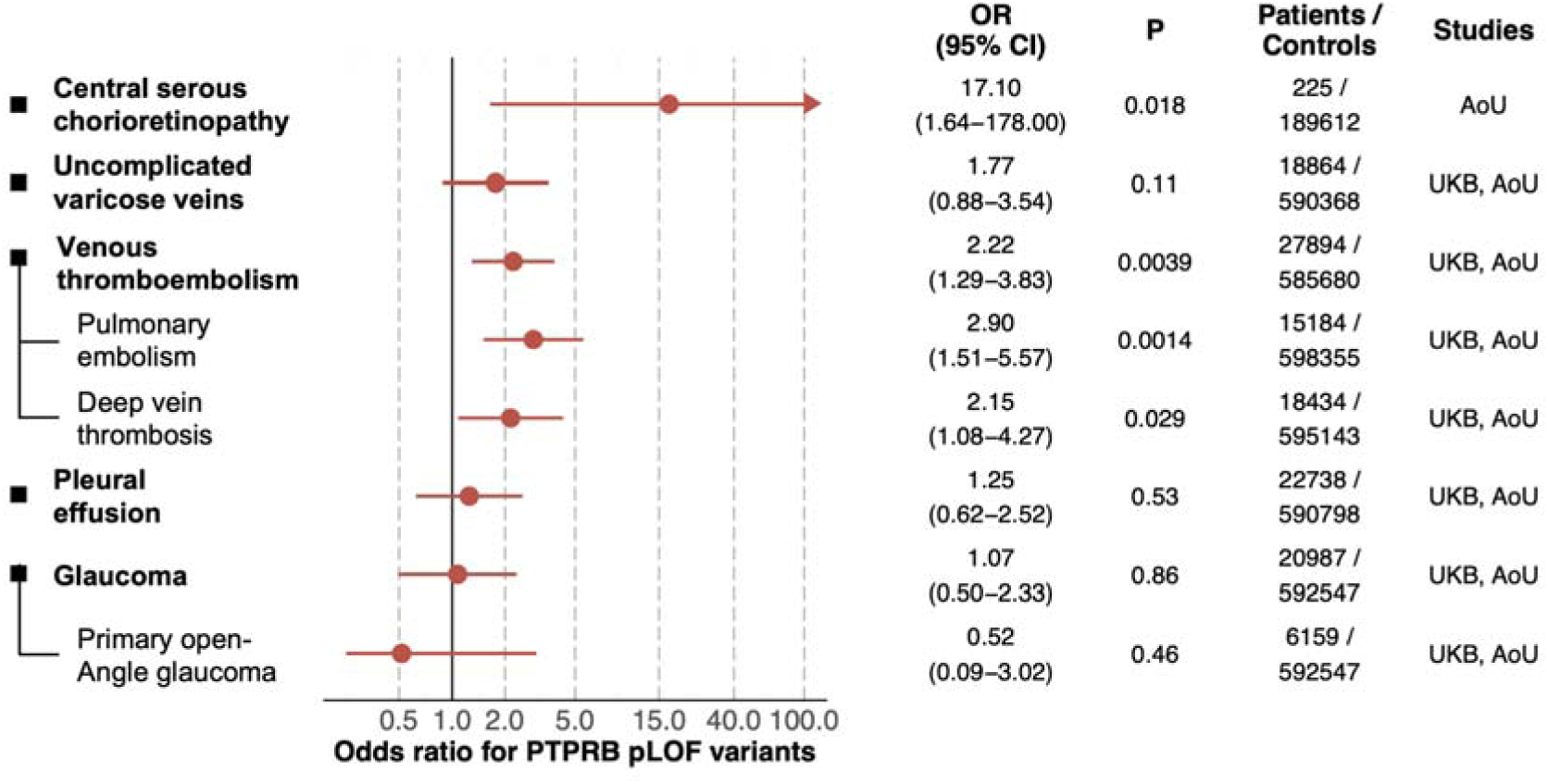
Disease associations of predicted loss-of-function variants in *PTPRB* among UK Biobank and All of Us participants. The risk of ocular and vascular disease of interest was evaluated for participants with a predicted loss-of-function (pLOF) variant in *PTPRB* in All of Us and UK Biobank. Due to lack of ophthalmological outpatient clinic data in UK Biobank participants, patients with central serous chorioretinopathy were only evaluated in All of Us. Examined diseases were selected based on the most significant associations observed for rs113791087 in FinnGen. Pulmonary embolism and deep vein thrombosis were examined separately as the major subtypes of venous thromboembolism, and primary open-angle glaucoma was examined separately as a major subtype of glaucoma. Association results from UK Biobank and All of Us were combined in an inverse variance weighted meta-analysis.

## Discussion

Despite progress in characterizing common variant genetic risk loci for CSC, pointing to specific genes and mechanisms that explain pathophysiology remains challenging due to the inherent hurdles faced when interpreting noncoding lead variants in GWAS. In contrast, here we identified a low-frequency missense variant (rs113791087) in *PTPRB,* the gene encoding VE-PTP, that was associated with a markedly increased risk of CSC and did not appear to be complicated by large stretches of linkage disequilibrium. Unexpectedly, rs113791087 was also associated with an increased risk of varicose veins and with a reduced risk of glaucoma. These previously uncharacterized genetic associations point directly to VE-PTP and demonstrate its relevance in ocular and systemic vascular diseases, support a role for vascular dysfunction in CSC, and in the future may inform therapeutic development.

VE-PTP is predominantly expressed as a membrane-bound protein in venous and arterial vascular endothelial cells where it functions as an important regulator of angiogenesis, vascular integrity, and vascular permeability^18–20^. Among its binding partners, the actions of VE-PTP via Tie-2 and VE-cadherin are best characterized. Through its intracellular domain, VE-PTP acts as a dephosphorylating inhibitor of the tyrosine kinase receptor Tie-2, an essential regulator of angiogenesis and vascular maintenance. VE-PTP additionally acts as a supporting regulator of VE-cadherin—a key adhesion molecule of adherens junctions between vascular endothelial cells^18,20,21^—potentially through both dephosphorylation and direct non-enzymatic transmembrane binding.

We are not aware of previous genome-wide significant associations between *PTPRB* variants and CSC, varicose veins, or glaucoma. A suggestive association with CSC—but not with varicose veins or glaucoma—has been previously reported for a different missense variant (rs61758735) in *PTPRB* based on an exome sequencing study of 2 families, and common intronic variants in the gene encoding VE-cadherin have been suggestively linked with CSC in patient cohorts^15,22^, but none of these reached genome-wide significance.

Somatic truncating variants in *PTPRB* have been previously linked with angiosarcoma^23^, and an exome-wide significant association between rare genetic variants in *PTPRB* in aggregate and intraocular pressure has been identified among UKB participants, although no direction of effect was reported^24^.

It is not known whether the disease associations of germline genetic variation in VE-PTP are predominantly mediated by changes in Tie-2 signaling, VE-cadherin, or unrelated mechanisms. At least partial relevance of Tie-2 signaling for glaucoma and venous dysfunction is suggested by the earlier identification of loss-of-function variants in the gene encoding Tie-2 in patients with primary congenital glaucoma^25^ and activating variants in patients with familial venous malformations with dilated vascular channels^26,27^. In mice, genetic insufficiency of Angiopoietin-1—a ligand of Tie-2—may inhibit normal formation of Shlemm’s canal and raise intraocular pressure^28^, genetic insufficiency of Tie-2 during embryogenesis may arrest venous development, and postnatal deletion of Tie-2 may lead to retinal venous degeneration with hemangioma-like vascular tufts^29^. Alternatively, or in addition to Tie-2 signaling, the disease associations observed for *PTPRB* variants might be mediated by reduced support for VE-cadherin, which is essential for normal blood vessel morphogenesis, mechanosensing, and permeability^30,31^.

The underlying cause of CSC is unknown, but an analogy between CSC and varicose veins has been drawn based on findings of delayed choroidal filling, venous dilation, and vascular hyperpermeability in choroidal angiography of patients with CSC, as highlighted by Spaide *et al*^8^. Statistical evidence exists for an overall enrichment of genes expressed in choroidal vascular endothelial cells among common variant CSC risk loci discovered to date^12^. Corticosteroid exposure—one of the classical triggers for CSC—may induce downregulation of VE-cadherin^22^, and VE-PTP may in general modulate vascular permeability in the context of increased inflammatory mediators^32–34^. These converging findings, together with the identification of a risk-increasing variant in VE-PTP in this study, provide support for a primary role of choroidal vascular incompetence in CSC. In contrast, other features that are commonly observed in patients with CSC such as disruption at the level of the RPE may represent downstream sequelae of vascular dysfunction^35^.

Genetic variants in VE-PTP may represent valuable instruments to guide therapeutic development for CSC. Modulation of Tie-2 signalling may prove to be a particularly attractive strategy, although the direction of modulation may differ depending on whether the genetic association with CSC is primarily mediated by Tie-2 or VE-Cadherin. If the genetic associations are mediated by reduced inhibition of Tie-2 by VE-PTP, *reducing* excessive Tie-2 activity could be favorable. Alternatively, if the genetic associations are due to reduced support of VE-PTP for VE-cadherin, it may be rather be possible to reduce VE-Cadherin internalization and rescue endothelial junction integrity by *increasing* Angiopoietin-1 or Tie-2 pathway activity^33,36–38^. Independent support for the potential benefit of increasing Angiopoietin-1 or Tie-2 activity comes from previous observations of low circulating levels of Angiopoietin-1, the primary agonist of Tie-2, in patients with both acute and chronic forms of CSC^39^, although such observational findings do not prove causality. Recently, inhibition of Angiopoietin-2—a natural antagonist of Angiopoietin-1 and Tie-2 activity^40^—has proved to be safe and effective in several vascular eye diseases including AMD, diabetic macular edema, and retinal vein occlusion^41–43^. Whether targeting Angiopoietin-2 could also be beneficial in patients with CSC is an uncertain but intriguing prospect.

In addition to the potential of this pathway in CSC, VE-PTP is also an existing investigational treatment target for other ocular diseases. Recent phase II clinical studies have evaluated razuprotafib (AKB-9778), an inhibitor of VE-PTP, as a novel treatment for glaucoma and diabetic macular edema, delivered through ophthalmic drops and subcutaneous injections, respectively^17,44–46^. The current findings and those of a recent exome sequencing study of UKB participants provide genetic support for VE-PTP as a therapeutic target for glaucoma^24^. The 18% reduction in the odds of glaucoma per minor allele of rs113791087 is greater than the effect estimate for any coding lead variant in a recent glaucoma GWAS meta-analysis and comparable with effect estimates reported for rare variants in *ANGPTL7*^47,48^, another recent genetically motivated therapeutic target. However, we note that the discordant effect directions for CSC and glaucoma warrant caution in therapeutic design to avoid unintended chorioretinal adverse effects with the use of VE-PTP inhibitors.

Lastly, inhibition of VE-PTP has been proposed as a potential treatment for selected high-grade cancers^49–51^. Germline genetic variants are not ideal instruments for complex oncological outcomes, and the lack of significant associations of rs113791087 with cancer endpoints in this study does not constitute strong evidence against the potential usefulness of VE-PTP inhibition. However, the increased risk of varicose veins and venous thromboembolism in participants with rs113791087 raises concern for unintended venous dysfunction from systemic VE-PTP inhibition, particularly as many patients with cancer are already at high risk of venous thromboembolism^52^.

Our findings should be interpreted in the context of study design. First, the ascertainment of CSC patients in most of the study datasets was based on ICD-10 codes. Reassuringly, significant associations for rs113791087 across different healthcare systems and in an outpatient clinic cohort based on expert clinician review support a true association with CSC. Second, the functional significance of rs113791087 is not known, and mechanistic studies are needed to understand its potential effects on the expression and activity of VE-PTP in the choroid. Directionally concordant although suggestive associations of *PTPRB* pLOF variants with CSC indicate that rs113791087 may act via a loss-of-function mechanism. Third, analyses of rs113791087 were limited to patients of genetically inferred European ancestry due to the low frequency of the variant in other populations; however, we included all participants from UKB and All of Us when assessing rare pLOF variants.

In summary, we identified a low-frequency missense variant in the gene encoding VE-PTP that was associated with a significantly increased risk of CSC and varicose veins and with a reduced risk of glaucoma. Our findings provide support for a central role of venous dysfunction in CSC and implicate the interconnected vascular endothelial function regulators VE-PTP, Tie-2 and VE-cadherin as potential therapeutic targets in diverse ocular and systemic vascular diseases.

## Data Availability

Individual-level genotypes and register data from FinnGen participants can be accessed by approved researchers via the Fingenious portal (https://site.fingenious.fi/en/) hosted by the Finnish Biobank Cooperative FinBB (https://finbb.fi/en/). Access to individual-level UKB data may be requested by researchers in academic, commercial, and charitable organizations. This study used data from the All of Us Research Program Controlled Tier Dataset v7, available to authorized users on the Researcher Workbench. Summary-level association data in MVP are available through dbGaP, with accession phs001672.v11.p1.

## Appendices

### Data availability

Individual-level genotypes and register data from FinnGen participants can be accessed by approved researchers via the Fingenious portal (https://site.fingenious.fi/en/) hosted by the Finnish Biobank Cooperative FinBB (https://finbb.fi/en/). Access to individual-level UKB data may be requested by researchers in academic, commercial, and charitable organizations. This study used data from the All of Us Research Program’s Controlled Tier Dataset v7, available to authorized users on the Researcher Workbench. Summary-level association data in MVP are available through dbGaP, with accession phs001672.v11.p1.

## Acknowledgements

We want to acknowledge the participants and investigators of FinnGen study. The FinnGen project is funded by two grants from Business Finland (HUS 4685/31/2016 and UH 4386/31/2016) and the following industry partners: AbbVie Inc., AstraZeneca UK Ltd, Biogen MA Inc., Bristol Myers Squibb (and Celgene Corporation & Celgene International II Sàrl), Genentech Inc., Merck Sharp & Dohme LCC, Pfizer Inc., GlaxoSmithKline Intellectual Property Development Ltd., Sanofi US Services Inc., Maze Therapeutics Inc., Janssen Biotech Inc, Novartis Pharma AG, and Boehringer Ingelheim International GmbH. Following biobanks are acknowledged for delivering biobank samples to FinnGen: Auria Biobank (www.auria.fi/biopankki), THL Biobank (www.thl.fi/biobank), Helsinki Biobank (www.helsinginbiopankki.fi), Biobank Borealis of Northern Finland (https://www.ppshp.fi/Tutkimus-ja-opetus/Biopankki/Pages/Biobank-Borealis-briefly-in-English.aspx), Finnish Clinical Biobank Tampere (www.tays.fi/en-US/Research_and_development/Finnish_Clinical_Biobank_Tampere), Biobank of Eastern Finland (www.ita-suomenbiopankki.fi/en), Central Finland Biobank (www.ksshp.fi/fi-FI/Potilaalle/Biopankki), Finnish Red Cross Blood Service Biobank (www.veripalvelu.fi/verenluovutus/biopankkitoiminta), Terveystalo Biobank (www.terveystalo.com/fi/Yritystietoa/Terveystalo-Biopankki/Biopankki/) and Arctic Biobank (https://www.oulu.fi/en/university/faculties-and-units/faculty-medicine/northern-finland-birth-cohorts-and-arctic-biobank). All Finnish Biobanks are members of BBMRI.fi infrastructure (www.bbmri.fi). Finnish Biobank Cooperative -FINBB (https://finbb.fi/) is the coordinator of BBMRI-ERIC operations in Finland.

We thank the U.S. veteran participants in MVP and MVP staff. This publication does not necessarily represent the views of the U.S. Department of Veterans Affairs or the United States Government. Support from the VA Office of Research & Development is acknowledged by N.S.P. (I01BX003364, I01 BX04557, IK6BX005233). We acknowledge the VA Million Veteran Program (MVP) and the VA-DOE genome-wide PheWAS core analytic team for generating the corresponding PheWAS summary statistics that were used in this manuscript.

The All of Us Research Program is supported by the National Institutes of Health, Office of the Director: Regional Medical Centers: 1 OT2 OD026549; 1 OT2 OD026554; 1 OT2 OD026557; 1 OT2 OD026556; 1 OT2 OD026550; 1 OT2 OD 026552; 1 OT2 OD026553; 1 OT2 OD026548; 1 OT2 OD026551; 1 OT2 OD026555; IAA #: AOD 16037; Federally Qualified Health Centers: HHSN 263201600085U; Data and Research Center: 5 U2C OD023196; Biobank: 1 U24 OD023121; The Participant Center: U24 OD023176; Participant Technology Systems Center: 1 U24 OD023163; Communications and Engagement: 3 OT2 OD023205; 3 OT2 OD023206; and Community Partners: 1 OT2 OD025277; 3 OT2 OD025315; 1 OT2 OD025337; 1 OT2 OD025276. In addition, the All of Us Research Program would not be possible without the partnership of its participants.

UK Biobank is generously supported by its founding funders the Wellcome Trust and UK Medical Research Council, as well as the British Heart Foundation, Cancer Research UK, Department of Health, Northwest Regional Development Agency and Scottish Government.

## Author Contributions

Conceptualization, E.J.R, M.J.D., S.K.I., P.T.E, C.JF.B., J.A.T., N.S.P., J.T.R. and B.R.G.; genomic analysis, J.T.R., B.R.G., L-C.W., S.J.J., X.W., SH.C., Y.L. and E.J.R., optical coherence tomography review, E.J.R., P.S., and M.G.T.; data collection and processing, E.H.C.vD., J.B., S.Y., C.J.F.B., A.P., M.J.D., C.W.H., S.K.I., J.T.A., K.K., W-C.W, M.B.G., C.L.N. and S.P.; funding acquisition, E.J.R., M.J.D., S.K.I., P.T.E., C.J.F.B., N.S.P. and S.Y.; supervision, E.J.R., M.J.D., S.K.I., P.T.E., C.JF.B., J.T.A and N.S.P; initial draft writing, J.T.R., E.J.R., B.R.G. and L-C.W.; review & editing, all authors.

## Online Methods

### Study design

FinnGen is a public-private partnership research project that combines genotype data from newly collected and legacy samples administered by Finnish biobanks (https://www.finngen.fi/en) to provide novel insight into human diseases. This study includes genotype data from 500,348 individuals from FinnGen Data Freeze 12. The data were linked by unique national personal identification numbers to the national hospital discharge registry (available from 1968) and the specialist outpatient registry (1998–).

The Million Veteran Program is a population-scale biobank within the Department of Veterans Affairs. Voluntary enrollment of veterans receiving care in the VA began in 2011^53^. All samples were scanned on the MVP 1.0 Axiom array (ThermoFisher); details on the design and QC are provided elsewhere^54^. Samples were phased using SHAPEIT4 and imputed to the TOPMed reference panel (version r2; N= 97,256 samples) using Minimac4. Phenotypes were based on electronic health record data with follow-up through the end of 2022.

The All of Us Research Program opened for enrollment in May 2018 and plans to enroll at least 1 million persons in the United States, collecting electronic health record data and biospecimens to advance the prevention and treatment of diseases^55^. In the current project, we used genetic data from 245,394 short-read whole genome-sequencing samples in the v7 release (**Supplementary Methods**), and excluded participants who did not have any electronic health record data.

Details for the European chronic CSC cohort have been reported previously^10^. For the genomic study, 521 European patients were recruited from the outpatient clinics of the Radboud University Medical Center (Netherlands), University Hospital of Cologne (Germany), and Leiden University Medical Center (Netherlands). Controls included 3,577 participants in the Nijmegen Biomedical Study. The patients with chronic CSC had subretinal fluid in at least one eye, retinal pigment epithelium irregularities with characteristic leakage on fluorescein angiography and corresponding hyperfluorescence on ICG.

UK Biobank (UKB) is a deeply phenotyped and genotyped prospective population-level cohort which recruited approximately 500,000 participants aged 40–69 in the UK between 2006–2010^56^. UKB participants were genotyped on the Affymetrix Applied Biosystems UK BiLEVE Axiom Array and the Affymetrix Applied Biosystems UKB Axiom Array. Sample and variant QC are described in detail by Bycroft et al^56^. rs113791087 was directly genotyped with high quality statistics (missingness rate of 0.00198). PCA was performed using fastPCA as described and the first 10 PCs were downloaded and used as covariates in further analysis^56^. OCT imaging was performed for a subset of UKB participants^57^. For this study, UKB data was accessed under applications #50211 and #17488.

More details for genotyping, imputation and sequencing in the different studies are described in the **Supplementary Methods**.

### Ethics statement

Study subjects in FinnGen provided informed consent for biobank research, based on the Finnish Biobank Act. Alternatively, separate research cohorts, collected between the Finnish Biobank Act coming into effect (in September 2013) and the start of FinnGen (August 2017), were collected as described in the **Supplementary Methods**.

The MVP024 study protocol was approved by the Veterans Affairs (VA) central Institutional Review Board (IRB). MVP participants provided written informed consent.

Use of All of Us data was approved under a data use agreement between the Massachusetts General Hospital and the All of Us research program. Study participants provided written informed consent.

The study of European patients with choronic CSC was carried out in accordance with the tenets of the Declaration of Helsinki and was approved by the local ethics committees of the Radboudumc, Leiden University Medical Center, and University Hospital of Cologne. Written informed consent was obtained for all participants.

The UKB OCT substudy was approved by the North West Multi-centre Research Ethics Committee in accordance with the principles of the Declaration of Helsinki. Written, informed consent was obtained for all UKB participants.

### Phenotype ascertainment

For the discovery GWAS, we identified patients with CSC based on the presence of at least one instance of the Finnish version of the *International Classification of Diseases 10th revision* (ICD-10) diagnosis code H35.7. Additionally, controls with retinal or choroidal disorders were excluded using the ICD-10 codes H30-H36 or any related endpoint as described previously (https://risteys.finregistry.fi/endpoints/H7_RETINASEPAR).

We conducted additional sensitivity analyses to evaluate bias due to potential confounders. In these analyses, instead of control-specific exclusion criteria, we applied the following exclusion criteria to both patients and controls: 1) at least one instance of an ICD-10 code corresponding to age-related macular degeneration (H35.30 or H35.31), and 2) at least one instance of any ICD-10 code corresponding to a larger set of potentially confounding causes of fluid maculopathy (**Supplementary Table 3**).

In MVP and All of Us, patients with CSC were identified based on at least one instance of the ICD-10-CM code H35.71* or ICD-9-CM code 362.41, and all participants with AMD (ICD-10: H35.1*, H35.2*, H35.3*; ICD-9: 362.5, 362.51, 362.52) were exclude. Case definitions for the European chronic CSC cohort have been described previously^10^. Ages of individuals in All of Us were calculated at 2022/7/1 or date of death.

### Genome-wide association study and regional meta-analysis of CSC

All FinnGen GWAS were conducted using Regenie v 2.2.4^58^, with sex, age at death or end of follow-up, principal components (PCs) 1–10, genotyping array, and genotyping batch as fixed-effect covariates. An approximate Firth correction was used for variants reaching nominal significance (*P*<0.01) in initial tests, and standard errors were computed based on the Firth beta estimate and Firth P-value.

Genomic analysis of CSC in MVP was performed using SAIGE v1.3.0^59^ on the set of samples classified as European ancestry using the HARE (Harmonized Ancestry and Race/Ethnicity) method^60^. We used leave-one-chromosome-out (LOCO) model fitting and enabled Firth effect size estimation for variants with *P*<0.05. Sex, age at enrollment, mean-centered age-squared, and the first ten within-ancestry PCs were included as covariates.

Genomic analysis of CSC in All of Us was performed using Regenie v3.2.2 with age at death or end of follow-up, (age at death or end of follow-up)^2^, sex, and PCs 1-5 and 15 (based on association with CSC at p < 0.05 in a separate association test).

To evaluate whether rs113791087 was the lead variant in its locus on chromosome 12 even when combining data from 4 different studies, we performed an inverse variance weighted meta-analysis of CSC for all genomic variants in the region of rs113791087 with GWAMA (v2.2.2). LocusZoom was used to generate locus plots for the meta-analysis using the European ancestry reference panel^61^.

More detailed methods for participating cohorts are described in the **Supplementary Information**.

### Analysis of Optical Coherence Tomography images in UKB

Deidentified OCT images of patients with the rs113791087 G allele (GG or GT) along with age-matched controls lacking the G allele (TT) were obtained from UKB (642 GG/GT OCTs, 1,058 TT OCTs; 10 age-matched participants with the TT genotype were pulled for every 1 participant with the GT or GG genotype, of which ∼16% had an OCT)^57^. Repeat OCTs from the same patient were removed. Images were randomly chosen and independently evaluated by 3 retina specialists who were blinded to genotype. Because the scope of manual OCT review has practical limitations, only the left eye of each patient underwent assessment and 708 OCTs were reviewed. Each grader was tasked with identifying and categorizing various types of RPE abnormalities (drusen, pattern dystrophy, pigment epithelial detachment or nonspecific retinal pigment epithelium irregularity, subretinal fluid, pachychoroid pigment epitheliopathy, atrophy, intraretinal fluid and/or evidence of CSC). Statistical significance was evaluated using linear regression in R including age, sex, examiner and the first 10 PCs as covariates.

### Phenome-wide association study, replication and meta-analyses

We evaluated the association of rs113791087 with 2,469 traits in FinnGen using Regenie v2.2.4 similarly to the association analyses for CSC.^58^ Phenotype definitions and the characteristics of study participants for these traits are publicly available on https://risteys.finregistry.fi/.

For top traits identified in this phenome-wide association study, we pursued single-variant replication and meta-analyses including data from FinnGen, UKB, MVP and All of Us. Where possible, custom disease definitions to match these outcomes were created in UKB (**Supplementary Table 8**), All of Us (**Supplementary Table 9**), and Million Veteran Program (**Supplementary Methods**).

We evaluated the association of rs113791087 with each outcome in unrelated participants of genetically inferred European ancestry (excluding second degree or closer relatives in UK Biobank and Million Veteran programs, and based on a kinship score cutoff of 0.1 in All of Us) with Firth logistic regression. Covariates in UK Biobank included genotyped sex, age, the first 10 PCs and genotyping array. Covariates in All of Us included age, age squared, sex, first 5 PCs, and additional outcome-related PCs (P<0.05 in separated models). Covariates for analyses of uncomplicated varicose veins in MVP included age, age^2^, sex and the first 10 PCs. For other phenotypes in MVP, we used European-ancestry (HARE) summary statistics generated by the genome-wide PheWAS (gwPheWAS) project^62^. Briefly, outcomes were derived from phecodes following standard definitions^63^, surveys distributed to all MVP enrollees^53^, and clinical laboratory and vital signs measurements. A linear or logistic regression GWAS was performed on each phenotype in a modified version of SAIGE using sex, age, age^2^, and the first 10 PCs as covariates.

Results from different studies were combined in an inverse variance weighted meta-analysis as implemented in the metagen() function of the *meta* package (v6.5-0) in R (v4.2.0).

We additionally evaluated the association of rs113791087 with the mean of the left and right eye corneal-compensated intraocular pressure (IOP) measurements in UKB participants. Participants with a history of surgery for glaucoma were excluded and IOP readings less than 5 or greater than 60 were excluded. Linear regression analyses were adjusted by genotyped sex, age, the first 10 PCs and genotyping array.

Lastly, we identified rare predicted loss-of-function (pLOF) variants in *PTPRB* among UKB and All of Us participants by using the LOFTEE algorithm, excluding low-confidence or flagged pLOF variants and variants present at a frequency above 0.1% in either biobank or across gnomAD v2 super-populations^64^. Associations with disease outcomes were evaluated with Firth logistic regression using similar analysis designs as for rs113791087, except all individuals were included regardless of genetically inferred ancestry.

### Additional epidemiological analyses

The association of CSC and varicose veins was evaluated in all available FinnGen participants using logistic regression as implemented in the *glm* function in R (v4.3.2) with CSC as the outcome and varicose veins, sex, and age at death or end of follow-up as independent predictors.

