## Supplementary Information for "Rare genetic variation in VE-PTP is associated with central serous chorioretinopathy, venous dysfunction and glaucoma"

### Supplementary Methods

#### FinnGen genotyping and imputation

Newly collected FinnGen samples were genotyped using a FinnGen ThermoFisher Axiom custom array (Thermo Fisher Scientific, San Diego, CA, USA), and legacy cohorts were genotyped using Illumina and Affymetrix arrays (Illumina Inc., San Diego, and Thermo Fisher Scientific, Santa Clara, CA, USA) as detailed previously[1]. Principal component analysis was used to remove samples who were not of genotype-inferred Finnish ancestry. Genotype imputation was performed using a population-specific SISu v4 imputation reference panel comprised of 8,557 whole genomes based on the protocol available at: https://dx.doi.org/10.17504/protocols.io.xbgfijw.

For rs113791087, genotype calls were based on genotyping in the majority of individuals and imputation with high quality (INFO score = 0.997) in the remaining individuals. Whole exome sequencing based calls in a subset of participants had good concordance with genotyping (**Supplementary Figure 3**).

#### FinnGen ethics statement

Study subjects in FinnGen provided informed consent for biobank research, based on the Finnish Biobank Act. Alternatively, separate research cohorts, collected prior the Finnish Biobank Act came into effect (in September 2013) and start of FinnGen (August 2017), were collected based on study-specific consents and later transferred to the Finnish biobanks after approval by Fimea (Finnish Medicines Agency), the National Supervisory Authority for Welfare and Health. Recruitment protocols followed the biobank protocols approved by Fimea. The Coordinating Ethics Committee of the Hospital District of Helsinki and Uusimaa (HUS) statement number for the FinnGen study is Nr HUS/990/2017.

The FinnGen study is approved by Finnish Institute for Health and Welfare (permit numbers: THL/2031/6.02.00/2017, THL/1101/5.05.00/2017, THL/341/6.02.00/2018, THL/2222/6.02.00/2018, THL/283/6.02.00/2019, THL/1721/5.05.00/2019 and THL/1524/5.05.00/2020), Digital and population data service agency (permit numbers: VRK43431/2017-3, VRK/6909/2018-3, VRK/4415/2019-3), the Social Insurance Institution (permit numbers: KELA 58/522/2017, KELA 131/522/2018, KELA 70/522/2019, KELA 98/522/2019, KELA 134/522/2019, KELA 138/522/2019, KELA 2/522/2020, KELA 16/522/2020), Findata permit numbers THL/2364/14.02/2020, THL/4055/14.06.00/2020, THL/3433/14.06.00/2020, THL/4432/14.06/2020, THL/5189/14.06/2020, THL/5894/14.06.00/2020, THL/6619/14.06.00/2020, THL/209/14.06.00/2021, THL/688/14.06.00/2021, THL/1284/14.06.00/2021, THL/1965/14.06.00/2021, THL/5546/14.02.00/2020, THL/2658/14.06.00/2021, THL/4235/14.06.00/2021, Statistics Finland (permit numbers: TK-53-1041-17 and TK/143/07.03.00/2020 (earlier TK-53-90-20) TK/1735/07.03.00/2021, TK/3112/07.03.00/2021) and Finnish Registry for Kidney Diseases permission/extract from the meeting minutes on 4th July 2019.

The Biobank Access Decisions for FinnGen samples and data utilized in FinnGen Data Freeze 11 include: THL Biobank BB2017_55, BB2017_111, BB2018_19, BB_2018_34, BB_2018_67, BB2018_71, BB2019_7, BB2019_8, BB2019_26, BB2020_1, BB2021_65, Finnish Red Cross Blood Service Biobank 7.12.2017, Helsinki Biobank HUS/359/2017, HUS/248/2020, HUS/430/2021 §28, §29, HUS/150/2022 §12, §13, §14, §15, §16, §17, §18, §23, §58 and §59, Auria Biobank AB17-5154 and amendment #1 (August 17 2020) and amendments BB_2021-0140, BB_2021-0156 (August 26 2021, Feb 2 2022), BB_2021-0169, BB_2021-0179, BB_2021-0161, AB20-5926 and amendment #1 (April 23 2020) and it´s modification (Sep 22 2021), BB_2022-0262, BB_2022-0256, Biobank Borealis of Northern Finland_2017_1013, 2021_5010, 2021_5018, 2021_5015, 2021_5015 Amendment, 2021_5023, 2021_5023 Amendment, 2021_5017, 2022_6001, 2022_6006 Amendment, BB22-0067, 2022_0262, Biobank of Eastern Finland 1186/2018 and amendment 22§/2020,

53§/2021, 13§/2022, 14§/2022, 15§/2022, 27§/2022, 28§/2022, 29§/2022, 33§/2022, 35§/2022, 36§/2022, 37§/2022, 39§/2022, 7§/2023, Finnish Clinical Biobank Tampere MH0004 and amendments (21.02.2020 & 06.10.2020), 8§/2021, 9§/2021, §9/2022, §10/2022, §12/2022, 13§/2022, §20/2022, §21/2022, §22/2022, §23/2022, 28§/2022, 29§/2022, 30§/2022, 31§/2022, 32§/2022, 38§/2022, 40§/2022, 42§/2022, 1§/2023, Central Finland Biobank 1-2017, BB_2021-0161, BB_2021-0169, BB_2021-0179, BB_2021-0170, BB_2022-0256, and Terveystalo Biobank STB 2018001 and amendment 25th Aug 2020, Finnish Hematological Registry and Clinical Biobank decision 18th June 2021, Arctic biobank P0844: ARC_2021_1001.

#### Genomic association analyses for additional custom outcomes in the Million Veteran Program

Participants with uncomplicated varicose veins were identified identified based on at least two instances of the ICD-10 codes I83.9* or ICD-9 code 454.9*. The association of rs113791087 with uncomplicated varicose veins was evaluated in participants of European ancestry who were unrelated within 2 degrees of relatedness and had a history of visiting eye clinics within the VA system. Independent predictors included rs113791087, age, $age^2$, sex and the first 10 genomic principal components.

Participants with dystrophies primarily involving the retinal pigment epithelium were identified in MVP using the ICD-9-CM code 362.76 and ICD-10-CM code 35.54. The association of rs113791087 with dystrophies primarily involving the retinal pigment epithelium was examined using logistic regression in participants of European ancestry who were unrelated within 2 degrees of relatedness and had a history of visiting eye clinics within the VA system. Independent predictors included rs113791087, age, $age^2$, sex and the first 10 genomic principal components.

#### Genomic data quality control and analyses in All of Us

The All of Us Research Program started enrollment in May 2018, planning to enroll over 1 million people from diverse groups in the United States[2]. Information collected includes health questionnaires, electronic health records (EHR), and physical measurements. Participants also provided biospecimens for genomic and other laboratory assessments.

In this current project, we used genetic data from 245,394 short-read whole genome-sequencing samples in the v7 release. We removed variants that 1) had GQ ≤ 20, 2) did not pass genotype filter (FT), 3) had call rate < 95%, or 4) were monomorphic. We kept individuals if they were 1) not included in the flag file (sample outlier QC failed), 2) European descent from genetic prediction, and 3) had EHR data available.

We used Regenie v3.2.2 to test the association of each variant in the *PTPRB* locus with CSC. In step 1, we used a smaller pruned dataset for model fitting and prediction. We performed two rounds of pruning in all individuals who passed variant QC by Plink, and 460,525 variants were included in our final pruned dataset (round 1: --indep-pairwise 100 5 0.1 --maf 0.005 --mac 100 --geno 0.01 --mind 0.01; round 2: --indep-pairwise 100 10 0.03). In step 2, we used the genomic predictions from step 1 to assess variant-outcome associations. Models were adjusted for age, age squared, sex, first 5 principal components (PC), and any additional outcome-associated PCs ($p<0.05$ in separate association tests with same covariate adjustment; PC15 for CSC). According to the All of Us data sharing policy, we excluded variants with estimated minor allele count < 40 from summary statistics.

Single-variant replication and analyses of predicted loss-of-function variants were carried out using Firth logistic regression. Related individuals were excluded based on the centrally provided table of related pairs (using a kinship score cutoff of 0.1), prioritising the inclusion of patients over references.

#### Exome sequencing and quality control in UK Biobank

Exomes were captured in UK Biobank using the revised version of the IDT xGen Exome Research Panel v1.0 on Illumina NovaSeq 6000 machines (https://www.ukbiobank.ac.uk/media/najcnoaz/access_064-uk-biobank-exome-release-faq_v11-1_final-002.pdf). Alignment using BWA-MEM, calling using DeepVariant, and joint genotyping using GLNexus have been described in detail elsewhere (https://biobank.ndph.ox.ac.uk/showcase/ukb/docs/UKB_WES_Protocol.pdf). For analyses of rare predicted loss-of-function variation in the present study, we used the OQFE exome call set and closely followed a previously published pipeline to perform stringent additional quality-control (QC), including genotype QC, variant QC and sample QC.[3] UK Biobank exome sequencing data analyses were performed under application number 17488 and were approved by the Mass General Brigham Institutional Review Board.

#### Predicted loss-of-function variant annotation in UK Biobank and All of Us

We annotated variants in UK Biobank and All of Us using the Loss-of-Function Transcript Effect Estimator (LOFTEE) plug-in implemented in the Variant Effect Predictor (VEP; v.105) (https://github.com/konradjk/loftee)[4]. Variants annotated as high-confidence loss-of-function variants by LOFTEE include frameshift indels, stop-gain variants and splice site disrupting variants. LOFs flagged for caution by LOFTEE were excluded. Variants were additionally annotated with the highest continental allele frequency based on gnomAD v2 exomes (including allele frequencies for each the European, East Asian, South Asian, African/African American and Latino/Admixed American super-populations).

### Supplementary Results

**Sources of diagnosis codes in FinnGen**

We evaluated the setting in which patients in FinnGen had received the ICD-10 diagnosis code H35.7. Among 1,477 patients, 1,471 (99.6%) had received at least one instance of the H35.7 diagnosis code in an ophthalmological specialty setting; 1048 (71.2%) of these patients had received at least one diagnosis code in a university hospital and 386 (26.2%) had received at least one diagnosis code in a central hospital within the Finnish healthcare system. Consistent with the classically self-resolving nature of CSC, only 45 (3.0%) of the patients had been included in the Finnish Register of Visual Impairment, which requires a corrected visual acuity permanently less than 0.3 in the better eye of the patient or a similar degree of permanent visual impairment.

#### Prevalence of potentially confounding diagnoses in FinnGen and All of Us

We evaluated the prevalence of potentially confounding diagnoses including diabetic retinopathy, retinal vein occlusion, pattern dystrophy, acute posterior multifocal placoid pigment epitheliopathy, myopic degeneration, toxic maculopathy, uveitis, Vogt-Koyanagi-Harada disease, inherited retinal disease or choroidal hemangioma among patients with CSC in FinnGen and All of Us. Any diagnosis of the aforementioned diseases was counted separately for patients with CSC and controls, counting both diagnoses before and after CSC diagnosis. Due to restrictions with reporting data from small sample numbers in the biobank-based studies, we were only able to evaluate these diagnoses in aggregate.

In total, 66 (6.0%) of patients with CSC and 16017 (3.3%) controls in FinnGen had any of the aforementioned diagnoses. In the Million Veteran Program, the aforementioned diagnoses were registered in fewer than 121 (17.1%) patients with CSC and 17363 (6.4%) controls. In

All of Us, the aforementioned diagnoses were registered in fewer than 20 (<15%) patients with CSC and 1522 (1.47%) controls. No CSC patients in the Million Veteran Program of All of Us had diagnoses of Vogt-Koyanagi-Harada disease; no precise ICD code for Vogt-Koyanagi-Harada was available in FinnGen.

### Supplementary Figures

#### Supplementary Figure 1. Quantile-quantile plot of the genome-wide association study of central serous chorioretinopathy in FinnGen

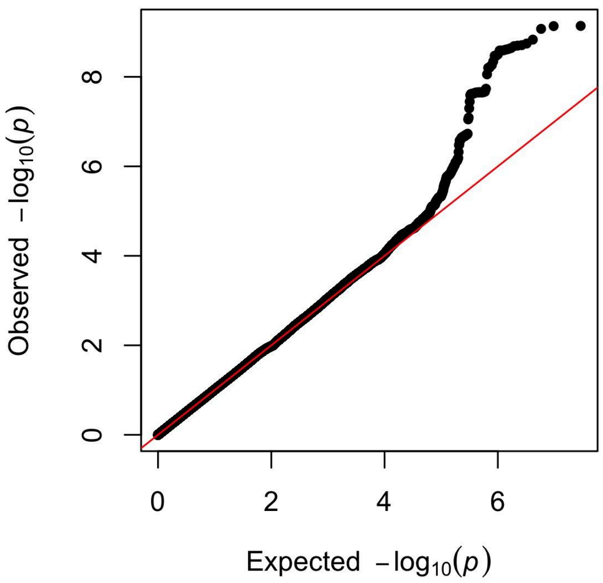

A genome-wide association study of central serous chorioretinopathy was conducted including 1,477 patients with central serous chorioretinopathy and 455,449 controls from the FinnGen study. The expected distribution (x-axis) and observed distribution (y-axis) of P-values is shown; the red line corresponds to the line of expectation. The genomic inflation factor was 1.018.

#### Supplementary Figure 2. Associations of variants in the *PTPRB* locus with central serous chorioretinopathy in FinnGen

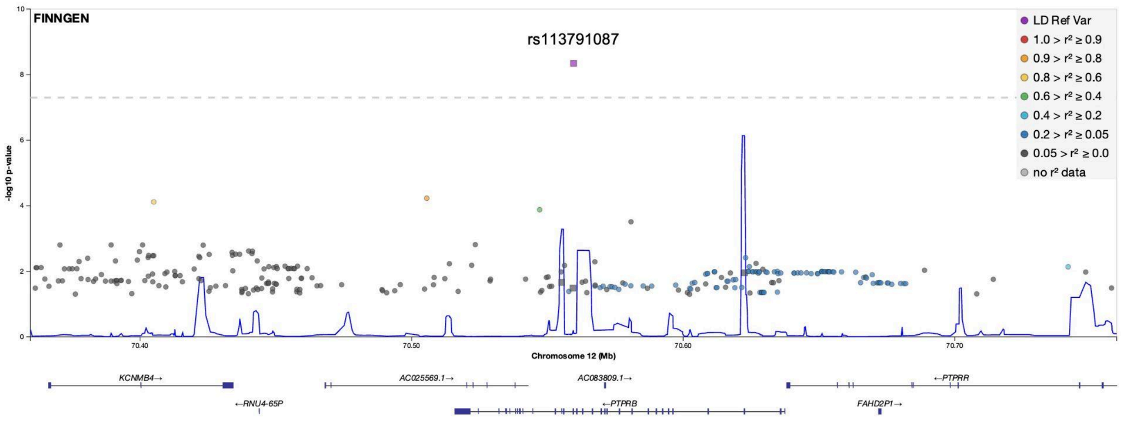

A genome-wide association study of central serous chorioretinopathy was conducted including 1,477 patients with central serous chorioretinopathy and 455,449 controls from the FinnGen study. Results are shown for the genome-wide significant locus containing the *PTPRB* gene on chromosome 12. Each genomic variant is plotted as a data point, with P-values shown on the y-axis on a logarithmic scale. Linkage disequilibrium between the lead variant (rs113791087, purple) and other variants in the region are shown on a colour scale.

Supplementary Figure 3. Genotype cluster plots for the *PTPRB* variant 12:70559589:T:G in FinnGen

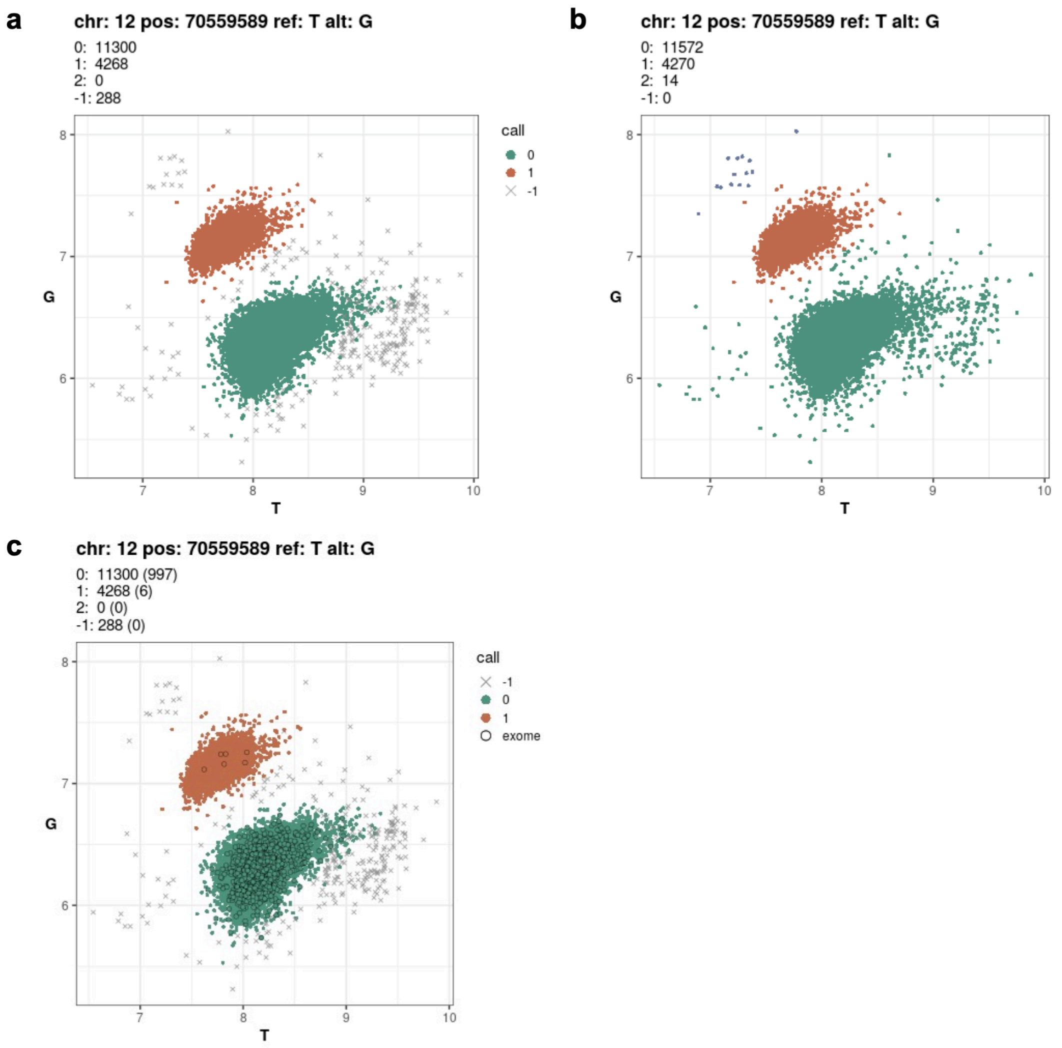

Panel **a** shows signal intensities and assigned genotype clusters based on chip data for 12:70559589:T:G (rs113791087) for 7,918 individuals from the FinnGen study. Panel **b** shows imputed genotypes for the same individuals overlaid on the signal intensities from chip data. Panel **c** shows exome sequencing calls in a subset of individuals demonstrating concordance with chip data.

Supplementary Figure 4. Associations of variants in the *PTPRB* locus with central serous chorioretinopathy in a meta-analysis of four study cohorts

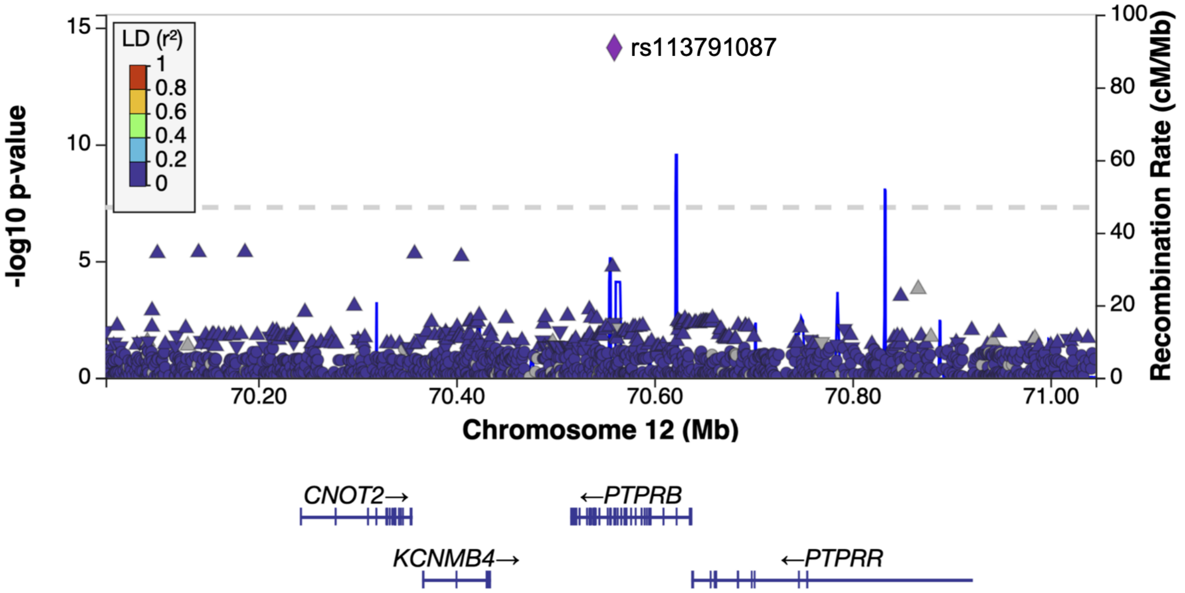

Variants in the chromosome 12 locus containing *PTPRB* were evaluated in a meta-analysis of genomic association studies of central serous chorioretinopathy. Data from a total of 2,452 patients and 881,210 controls from FinnGen, Million Veteran Program, All of Us, and a European chronic CSC cohort were included in the meta-analysis. Each genomic variant is plotted as a data point, with P-values shown on the y-axis on a logarithmic scale. The genome-wide significance threshold (P = 5e-8) is shown with a dashed line. Linkage disequilibrium (LD) between the lead variant (rs113791087) and other variants in the loci are shown on a color scale. The figure was generated with LocusZoom using the European-ancestry reference panel.

Supplementary Figure 5. Associations of variants in the *PTPRB* locus with central serous chorioretinopathy in All of Us

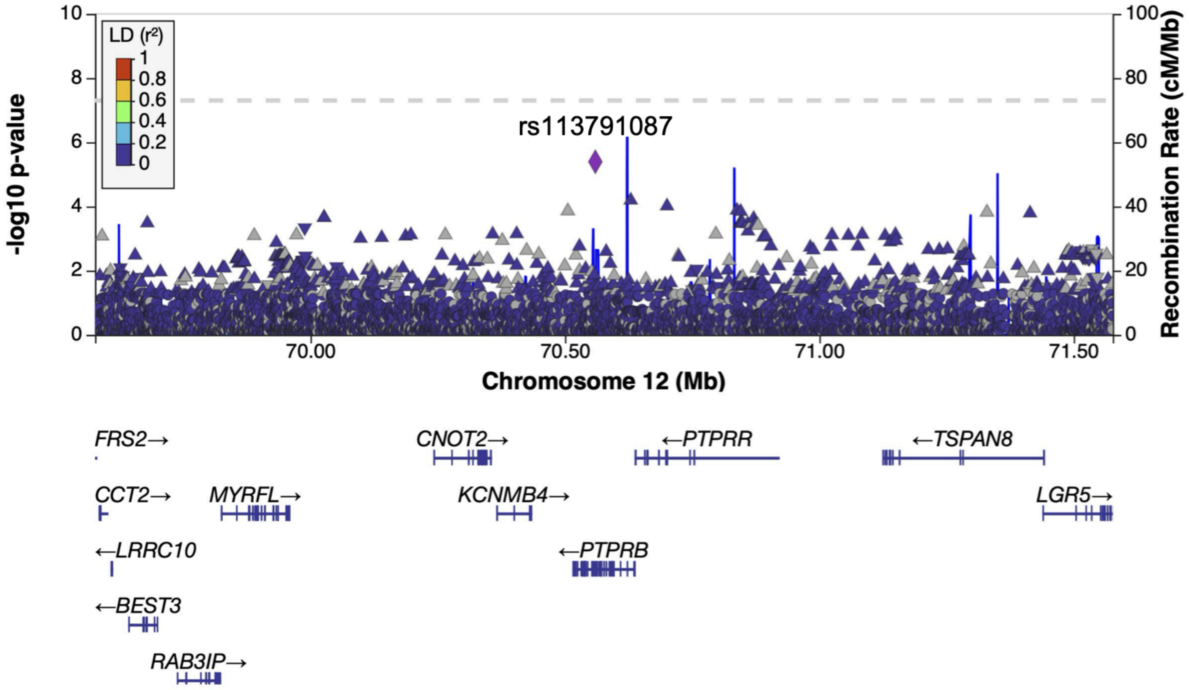

A case-control genomic association study was performed in All of Us, including 133 CSC patients and 119043 controls. Association results are shown for the region of the *PTPRB* gene, including all variants with a minor allele count over 40.

Supplementary Figure 6. Associations of variants in the *PTPRB* locus with varicose veins in FinnGen

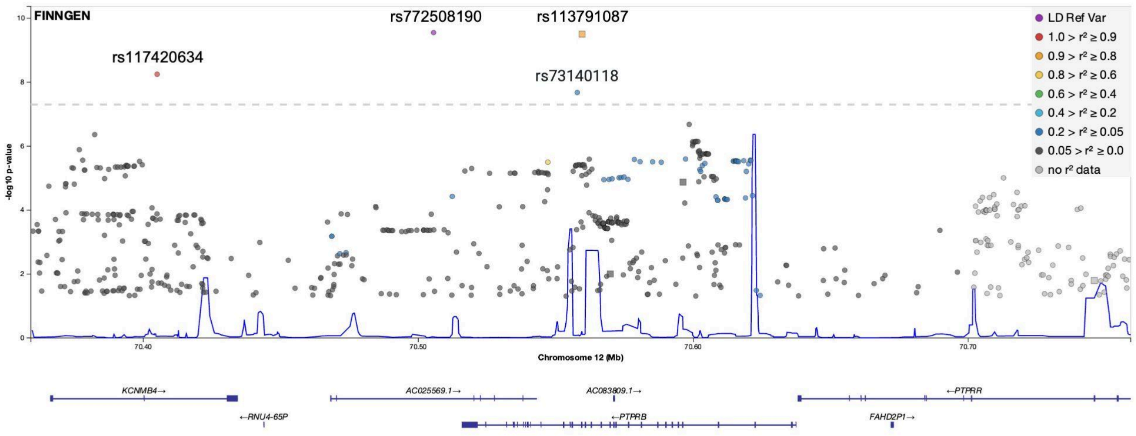

A genome-wide association study of varicose veins was conducted including 38,467 patients with varicose veins and 432,223 controls from the FinnGen study. Results are shown for the genome-wide significant locus containing the *PTPRB* gene on chromosome 12. Each genomic variant is plotted as a data point, with P-values shown on the y-axis on a logarithmic scale. Linkage disequilibrium between the lead variant (rs772508190, purple) and other variants in the region are shown on a colour scale.

### FinnGen Study Members

| Full Name | Affiliation | Role 1 | Role 2 |
|---|---|---|---|
| Aarno Palotie | Institute for Molecular Medicine Finland (FIMM), HiLIFE, University of Helsinki, Helsinki, Finland; Broad Institute of MIT and Harvard; Massachusetts General Hospital | Steering Committee | Steering Committee |
| Mark Daly | Institute for Molecular Medicine Finland (FIMM), HiLIFE, University of Helsinki, Helsinki, Finland; Broad Institute of MIT and Harvard; Massachusetts General Hospital | Steering Committee | Steering Committee |
| Bridget Riley-Gills | Abbvie, Chicago, IL, United States | Steering Committee | **Pharmaceutical companies** |
| Howard Jacob | Abbvie, Chicago, IL, United States | Steering Committee | **Pharmaceutical companies** |
| Coralie Viollet | Astra Zeneca, Cambridge, United Kingdom | Steering Committee | **Pharmaceutical companies** |
| Slavé Petrovski | Astra Zeneca, Cambridge, United Kingdom | Steering Committee | **Pharmaceutical companies** |
| Chia-Yen Chen | Biogen, Cambridge, MA, United States | Steering Committee | **Pharmaceutical companies** |
| Sally John | Biogen, Cambridge, MA, United States | Steering Committee | **Pharmaceutical companies** |
| George Okafo | Boehringer Ingelheim, Ingelheim am Rhein, Germany | Steering Committee | **Pharmaceutical companies** |
| Robert Plenge | Bristol Myers Squibb, New York, NY, United States | Steering Committee | **Pharmaceutical companies** |
| Joseph Maranville | Bristol Myers Squibb, New York, NY, United States | Steering Committee | **Pharmaceutical companies** |
| Mark McCarthy | Genentech, San Francisco, CA, United States | Steering Committee | **Pharmaceutical companies** |
| Rion Pendergrass | Genentech, San Francisco, CA, United States | Steering Committee | **Pharmaceutical companies** |
| Margaret G. Ehm | GlaxoSmithKline, Collegeville, PA, United States | Steering Committee | **Pharmaceutical companies** |
| Kirsi Auro | GlaxoSmithKline, Espoo, Finland | Steering Committee | **Pharmaceutical companies** |
| Simonne Longerich | Merck, Kenilworth, NJ, United States | Steering Committee | **Pharmaceutical companies** |
| Anders Mälarstig | Pfizer, New York, NY, United States | Steering Committee | **Pharmaceutical companies** |
| Anna Vlahiotis | Pfizer, New York, NY, United States | Steering Committee | **Pharmaceutical companies** |
| Katherine Klinger | Translational Sciences, Sanofi R&D, Framingham, MA, USA | Steering Committee | **Pharmaceutical companies** |
| Clement Chatelain | Translational Sciences, Sanofi R&D, Framingham, MA, USA | Steering Committee | **Pharmaceutical companies** |
| Matthias Gossel | Translational Sciences, Sanofi R&D, Framingham, MA, USA | Steering Committee | **Pharmaceutical companies** |
| Karol Estrada | Maze Therapeutics, San Francisco, CA, United States | Steering Committee | **Pharmaceutical companies** |
| Robert Graham | Maze Therapeutics, San Francisco, CA, United States | Steering Committee | **Pharmaceutical companies** |
| Dawn Waterworth | Janssen Research & Development, LLC, Spring House, PA, United States | Steering Committee | **Pharmaceutical companies** |
| Chris O´Donnell | Novartis Institutes for BioMedical Research, Cambridge, MA, United States | Steering Committee | **Pharmaceutical companies** |
| Nicole Renaud | Novartis Institutes for BioMedical Research, Cambridge, MA, United States | Steering Committee | **Pharmaceutical companies** |
| Tomi P. Mäkelä | HiLIFE, University of Helsinki, Finland, Finland | Steering Committee | **University of Helsinki & Biobanks** |

| | | | |
|---|---|---|---|
| Jaakko Kaprio | Institute for Molecular Medicine Finland (FIMM), HiLIFE, University of Helsinki, Helsinki, Finland | **Steering Committee** | **University of Helsinki & Biobanks** |
| Minna Ruddock | Arctic biobank / University of Oulu | **Steering Committee** | **University of Helsinki & Biobanks** |
| Petri Virolainen | Auria Biobank / University of Turku / Hospital District of Southwest Finland, Turku, Finland | **Steering Committee** | **University of Helsinki & Biobanks** |
| Antti Hakanen | Auria Biobank / University of Turku / Hospital District of Southwest Finland, Turku, Finland | **Steering Committee** | **University of Helsinki & Biobanks** |
| Terhi Kilpi | THL Biobank / Finnish Institute for Health and Welfare (THL), Helsinki, Finland | **Steering Committee** | **University of Helsinki & Biobanks** |
| Markus Perola | THL Biobank / Finnish Institute for Health and Welfare (THL), Helsinki, Finland | **Steering Committee** | **University of Helsinki & Biobanks** |
| Jukka Partanen | Finnish Red Cross Blood Service / Finnish Hematology Registry and Clinical Biobank, Helsinki, Finland | **Steering Committee** | **University of Helsinki & Biobanks** |
| Taneli Raivio | Helsinki Biobank / Helsinki University and Hospital District of Helsinki and Uusimaa, Helsinki | **Steering Committee** | **University of Helsinki & Biobanks** |
| Jani Tikkanen | Northern Finland Biobank Borealis / University of Oulu / Northern Ostrobothnia Hospital District, Oulu, Finland | **Steering Committee** | **University of Helsinki & Biobanks** |
| Raisa Serpi | Northern Finland Biobank Borealis / University of Oulu / Northern Ostrobothnia Hospital District, Oulu, Finland | **Steering Committee** | **University of Helsinki & Biobanks** |
| Kati Kristiansson | Finnish Clinical Biobank Tampere / University of Tampere / Pirkanmaa Hospital District, Tampere, Finland | **Steering Committee** | **University of Helsinki & Biobanks** |
| Veli-Matti Kosma | Biobank of Eastern Finland / University of Eastern Finland / Northern Savo Hospital District, Kuopio, Finland | **Steering Committee** | **University of Helsinki & Biobanks** |
| Jari Laukkanen | Central Finland Biobank / University of Jyväskylä / Central Finland Health Care District, Jyväskylä, Finland | **Steering Committee** | **University of Helsinki & Biobanks** |
| Marco Hautalahti | FINBB - Finnish biobank cooperative | **Steering Committee** | **University of Helsinki & Biobanks** |
| Outi Tuovila | Business Finland, Helsinki, Finland | **Steering Committee** | **Other Experts/ Non-Voting Members** |
| Jeffrey Waring | Abbvie, Chicago, IL, United States | **Scientific Committee** | **Pharmaceutical companies** |
| Bridget Riley-Gillis | Abbvie, Chicago, IL, United States | **Scientific Committee** | **Pharmaceutical companies** |
| Fedik Rahimov | Abbvie, Chicago, IL, United States | **Scientific Committee** | **Pharmaceutical companies** |
| Ioanna Tachmazidou | Astra Zeneca, Cambridge, United Kingdom | **Scientific Committee** | **Pharmaceutical companies** |
| Chia-Yen Chen | Biogen, Cambridge, MA, United States | **Scientific Committee** | **Pharmaceutical companies** |
| Zhihao Ding | Boehringer Ingelheim, Ingelheim am Rhein, Germany | **Scientific Committee** | **Pharmaceutical companies** |
| Marc Jung | Boehringer Ingelheim, Ingelheim am Rhein, Germany | **Scientific Committee** | **Pharmaceutical companies** |
| Hanati Tuoken | Boehringer Ingelheim, Ingelheim am Rhein, Germany | **Scientific Committee** | **Pharmaceutical companies** |
| Shameek Biswas | Bristol Myers Squibb, New York, NY, United States | **Scientific Committee** | **Pharmaceutical companies** |

| | | | |
|---|---|---|---|
| Rion Pendergrass | Genentech, San Francisco, CA, United States | Scientific Committee | **Pharmaceutical companies** |
| Margaret G. Ehm | GlaxoSmithKline, Collegeville, PA, United States | Scientific Committee | **Pharmaceutical companies** |
| David Pulford | GlaxoSmithKline, Stevenage, United Kingdom | Scientific Committee | **Pharmaceutical companies** |
| Neha Raghavan | Merck, Kenilworth, NJ, United States | Scientific Committee | **Pharmaceutical companies** |
| Adriana Huertas-Vazquez | Merck, Kenilworth, NJ, United States | Scientific Committee | **Pharmaceutical companies** |
| Jae-Hoon Sul | Merck, Kenilworth, NJ, United States | Scientific Committee | **Pharmaceutical companies** |
| Anders Mälarstig | Pfizer, New York, NY, United States | Scientific Committee | **Pharmaceutical companies** |
| Xinli Hu | Pfizer, New York, NY, United States | Scientific Committee | **Pharmaceutical companies** |
| Åsa Hedman | Pfizer, New York, NY, United States | Scientific Committee | **Pharmaceutical companies** |
| Katherine Klinger | Translational Sciences, Sanofi R&D, Framingham, MA, USA | Scientific Committee | **Pharmaceutical companies** |
| Robert Graham | Maze Therapeutics, San Francisco, CA, United States | Scientific Committee | **Pharmaceutical companies** |
| Dawn Waterworth | Janssen Research & Development, LLC, Spring House, PA, United States | Scientific Committee | **Pharmaceutical companies** |
| Nicole Renaud | Novartis Institutes for BioMedical Research, Cambridge, MA, United States | Scientific Committee | **Pharmaceutical companies** |
| Ma´en Obeidat | Novartis Institutes for BioMedical Research, Cambridge, MA, United States | Scientific Committee | **Pharmaceutical companies** |
| Jonathan Chung | Novartis Institutes for BioMedical Research, Cambridge, MA, United States | Scientific Committee | **Pharmaceutical companies** |
| Jonas Zierer | Novartis Institutes for BioMedical Research, Cambridge, MA, United States | Scientific Committee | **Pharmaceutical companies** |
| Mari Niemi | Novartis Institutes for BioMedical Research, Cambridge, MA, United States | Scientific Committee | **Pharmaceutical companies** |
| Samuli Ripatti | Institute for Molecular Medicine Finland (FIMM), HiLIFE, University of Helsinki, Helsinki, Finland | Scientific Committee | **University of Helsinki & Biobanks** |
| Johanna Schleutker | Auria Biobank / Univ. of Turku / Hospital District of Southwest Finland, Turku, Finland | Scientific Committee | **University of Helsinki & Biobanks** |
| Markus Perola | THL Biobank / Finnish Institute for Health and Welfare (THL), Helsinki, Finland | Scientific Committee | **University of Helsinki & Biobanks** |
| Mikko Arvas | Finnish Red Cross Blood Service / Finnish Hematology Registry and Clinical Biobank, Helsinki, Finland | Scientific Committee | **University of Helsinki & Biobanks** |
| Olli Carpén | Helsinki Biobank / Helsinki University and Hospital District of Helsinki and Uusimaa, Helsinki | Scientific Committee | **University of Helsinki & Biobanks** |
| Reetta Hinttala | Northern Finland Biobank Borealis / University of Oulu / Northern Ostrobothnia Hospital District, Oulu, Finland | Scientific Committee | **University of Helsinki & Biobanks** |
| Johannes Kettunen | Northern Finland Biobank Borealis / University of Oulu / Northern Ostrobothnia Hospital District, Oulu, Finland | Scientific Committee | **University of Helsinki & Biobanks** |
| Arto Mannermaa | Biobank of Eastern Finland / University of Eastern Finland / Northern Savo Hospital District, Kuopio, Finland | Scientific Committee | **University of Helsinki & Biobanks** |
| Katriina Aalto-Setälä | Faculty of Medicine and Health Technology, Tampere University, Tampere, Finland | Scientific Committee | **University of Helsinki & Biobanks** |
| Mika Kähönen | Finnish Clinical Biobank Tampere / University of Tampere / Pirkanmaa | Scientific | **University of** |

| | | | |
|---|---|---|---|
| | Hospital District, Tampere, Finland | Committee | **Helsinki & Biobanks** |
| Jari Laukkanen | Central Finland Biobank / University of Jyväskylä / Central Finland Health Care District, Jyväskylä, Finland | Scientific Committee | **University of Helsinki & Biobanks** |
| Johanna Mäkelä | FINBB - Finnish biobank cooperative | Scientific Committee | **University of Helsinki & Biobanks** |
| Reetta Kälviäinen | Northern Savo Hospital District, Kuopio, Finland | Clinical Groups | **Neurology Group** |
| Valtteri Julkunen | Northern Savo Hospital District, Kuopio, Finland | Clinical Groups | **Neurology Group** |
| Hilkka Soininen | Northern Savo Hospital District, Kuopio, Finland | Clinical Groups | **Neurology Group** |
| Anne Remes | Northern Ostrobothnia Hospital District, Oulu, Finland | Clinical Groups | **Neurology Group** |
| Mikko Hiltunen | University of Eastern Finland, Kuopio, Finland | Clinical Groups | **Neurology Group** |
| Jukka Peltola | Pirkanmaa Hospital District, Tampere, Finland | Clinical Groups | **Neurology Group** |
| Minna Raivio | Hospital District of Helsinki and Uusimaa, Helsinki, Finland | Clinical Groups | **Neurology Group** |
| Pentti Tienari | Hospital District of Helsinki and Uusimaa, Helsinki, Finland | Clinical Groups | **Neurology Group** |
| Juha Rinne | Hospital District of Southwest Finland, Turku, Finland | Clinical Groups | **Neurology Group** |
| Roosa Kallionpää | Hospital District of Southwest Finland, Turku, Finland | Clinical Groups | **Neurology Group** |
| Juulia Partanen | Institute for Molecular Medicine Finland, HiLIFE, University of Helsinki, Finland | Clinical Groups | **Neurology Group** |
| Adam Ziemann | Abbvie, Chicago, IL, United States | Clinical Groups | **Neurology Group** |
| Nizar Smaoui | Abbvie, Chicago, IL, United States | Clinical Groups | **Neurology Group** |
| Anne Lehtonen | Abbvie, Chicago, IL, United States | Clinical Groups | **Neurology Group** |
| Susan Eaton | Biogen, Cambridge, MA, United States | Clinical Groups | **Neurology Group** |
| Heiko Runz | Biogen, Cambridge, MA, United States | Clinical Groups | **Neurology Group** |
| Sanni Lahdenperä | Biogen, Cambridge, MA, United States | Clinical Groups | **Neurology Group** |
| Shameek Biswas | Bristol Myers Squibb, New York, NY, United States | Clinical Groups | **Neurology Group** |
| Natalie Bowers | Genentech, San Francisco, CA, United States | Clinical Groups | **Neurology Group** |
| Edmond Teng | Genentech, San Francisco, CA, United States | Clinical Groups | **Neurology Group** |
| Rion Pendergrass | Genentech, San Francisco, CA, United States | Clinical Groups | **Neurology Group** |
| Fanli Xu | GlaxoSmithKline, Brentford, United Kingdom | Clinical Groups | **Neurology Group** |
| David Pulford | GlaxoSmithKline, Stevenage, United Kingdom | Clinical Groups | **Neurology Group** |
| Kirsi Auro | GlaxoSmithKline, Espoo, Finland | Clinical Groups | **Neurology Group** |
| Laura Addis | GlaxoSmithKline, Brentford, United Kingdom | Clinical Groups | **Neurology Group** |
| John Eicher | GlaxoSmithKline, Brentford, United Kingdom | Clinical Groups | **Neurology Group** |
| Qingqin S Li | Janssen Research & Development, LLC, Titusville, NJ 08560, United States | Clinical Groups | **Neurology Group** |
| Karen He | Janssen Research & Development, LLC, Spring House, PA, United States | Clinical | **Neurology Group** |

| | | | |
|---|---|---|---|
| | | Groups | |
| Ekaterina Khramtsova | Janssen Research & Development, LLC, Spring House, PA, United States | Clinical Groups | **Neurology Group** |
| Neha Raghavan | Merck, Kenilworth, NJ, United States | Clinical Groups | **Neurology Group** |
| Martti Färkkilä | Hospital District of Helsinki and Uusimaa, Helsinki, Finland | Clinical Groups | **Gastroenterology Group** |
| Jukka Koskela | Hospital District of Helsinki and Uusimaa, Helsinki, Finland | Clinical Groups | **Gastroenterology Group** |
| Sampsa Pikkarainen | Hospital District of Helsinki and Uusimaa, Helsinki, Finland | Clinical Groups | **Gastroenterology Group** |
| Airi Jussila | Pirkanmaa Hospital District, Tampere, Finland | Clinical Groups | **Gastroenterology Group** |
| Katri Kaukinen | Pirkanmaa Hospital District, Tampere, Finland | Clinical Groups | **Gastroenterology Group** |
| Timo Blomster | Northern Ostrobothnia Hospital District, Oulu, Finland | Clinical Groups | **Gastroenterology Group** |
| Mikko Kiviniemi | Northern Savo Hospital District, Kuopio, Finland | Clinical Groups | **Gastroenterology Group** |
| Markku Voutilainen | Hospital District of Southwest Finland, Turku, Finland | Clinical Groups | **Gastroenterology Group** |
| Mark Daly | Institute for Molecular Medicine, Finland (FIMM), HiLIFE, University of Helsinki, Helsinki, Finland; Broad Institute of MIT and Harvard; Massachusetts General Hospital | Clinical Groups | **Gastroenterology Group** |
| Jeffrey Waring | Abbvie, Chicago, IL, United States | Clinical Groups | **Gastroenterology Group** |
| Nizar Smaoui | Abbvie, Chicago, IL, United States | Clinical Groups | **Gastroenterology Group** |
| Fedik Rahimov | Abbvie, Chicago, IL, United States | Clinical Groups | **Gastroenterology Group** |
| Anne Lehtonen | Abbvie, Chicago, IL, United States | Clinical Groups | **Gastroenterology Group** |
| Tim Lu | Genentech, San Francisco, CA, United States | Clinical Groups | **Gastroenterology Group** |
| Natalie Bowers | Genentech, San Francisco, CA, United States | Clinical Groups | **Gastroenterology Group** |
| Rion Pendergrass | Genentech, San Francisco, CA, United States | Clinical Groups | **Gastroenterology Group** |
| Linda McCarthy | GlaxoSmithKline, Brentford, United Kingdom | Clinical Groups | **Gastroenterology Group** |
| Amy Hart | Janssen Research & Development, LLC, Spring House, PA, United States | Clinical Groups | **Gastroenterology Group** |
| Meijian Guan | Janssen Research & Development, LLC, Spring House, PA, United States | Clinical Groups | **Gastroenterology Group** |
| Jason Miller | Merck, Kenilworth, NJ, United States | Clinical Groups | **Gastroenterology Group** |
| Kirsi Kalpala | Pfizer, New York, NY, United States | Clinical Groups | **Gastroenterology Group** |
| Melissa Miller | Pfizer, New York, NY, United States | Clinical Groups | **Gastroenterology Group** |
| Xinli Hu | Pfizer, New York, NY, United States | Clinical Groups | **Gastroenterology Group** |
| Kari Eklund | Hospital District of Helsinki and Uusimaa, Helsinki, Finland | Clinical Groups | **Rheumatology Group** |
| Antti Palomäki | Hospital District of Southwest Finland, Turku, Finland | Clinical Groups | **Rheumatology Group** |
| Pia Isomäki | Pirkanmaa Hospital District, Tampere, Finland | Clinical Groups | **Rheumatology Group** |
| Laura Pirilä | Hospital District of Southwest Finland, Turku, Finland | Clinical Groups | **Rheumatology Group** |
| Oili Kaipiainen-Seppänen | Northern Savo Hospital District, Kuopio, Finland | Clinical Groups | **Rheumatology Group** |

| | | | |
|---|---|---|---|
| Johanna Huhtakangas | Northern Ostrobothnia Hospital District, Oulu, Finland | Clinical Groups | **Rheumatology Group** |
| Nina Mars | Institute for Molecular Medicine Finland (FIMM), HiLIFE, University of Helsinki, Helsinki, Finland | Clinical Groups | **Rheumatology Group** |
| Jeffrey Waring | Abbvie, Chicago, IL, United States | Clinical Groups | **Rheumatology Group** |
| Fedik Rahimov | Abbvie, Chicago, IL, United States | Clinical Groups | **Rheumatology Group** |
| Apinya Lertratanakul | Abbvie, Chicago, IL, United States | Clinical Groups | **Rheumatology Group** |
| Nizar Smaoui | Abbvie, Chicago, IL, United States | Clinical Groups | **Rheumatology Group** |
| Anne Lehtonen | Abbvie, Chicago, IL, United States | Clinical Groups | **Rheumatology Group** |
| Coralie Viollet | AstraZeneca, Cambridge, United Kingdom | Clinical Groups | **Rheumatology Group** |
| Marla Hochfeld | Bristol Myers Squibb, New York, NY, United States | Clinical Groups | **Rheumatology Group** |
| Natalie Bowers | Genentech, San Francisco, CA, United States | Clinical Groups | **Rheumatology Group** |
| Rion Pendergrass | Genentech, San Francisco, CA, United States | Clinical Groups | **Rheumatology Group** |
| Jorge Esparza Gordillo | GlaxoSmithKline, Brentford, United Kingdom | Clinical Groups | **Rheumatology Group** |
| Kirsi Auro | GlaxoSmithKline, Espoo, Finland | Clinical Groups | **Rheumatology Group** |
| Dawn Waterworth | Janssen Research & Development, LLC, Spring House, PA, United States | Clinical Groups | **Rheumatology Group** |
| Fabiana Farias | Merck, Kenilworth, NJ, United States | Clinical Groups | **Rheumatology Group** |
| Kirsi Kalpala | Pfizer, New York, NY, United States | Clinical Groups | **Rheumatology Group** |
| Nan Bing | Pfizer, New York, NY, United States | Clinical Groups | **Rheumatology Group** |
| Xinli Hu | Pfizer, New York, NY, United States | Clinical Groups | **Rheumatology Group** |
| Tarja Laitinen | Pirkanmaa Hospital District, Tampere, Finland | Clinical Groups | **Pulmonology Group** |
| Margit Pelkonen | Northern Savo Hospital District, Kuopio, Finland | Clinical Groups | **Pulmonology Group** |
| Paula Kauppi | Hospital District of Helsinki and Uusimaa, Helsinki, Finland | Clinical Groups | **Pulmonology Group** |
| Hannu Kankaanranta | University of Gothenburg, Gothenburg, Sweden/ Seinäjoki Central Hospital, Seinäjoki, Finland/ Tampere University, Tampere, Finland | Clinical Groups | **Pulmonology Group** |
| Terttu Harju | Northern Ostrobothnia Hospital District, Oulu, Finland | Clinical Groups | **Pulmonology Group** |
| Riitta Lahesmaa | Hospital District of Southwest Finland, Turku, Finland | Clinical Groups | **Pulmonology Group** |
| Nizar Smaoui | Abbvie, Chicago, IL, United States | Clinical Groups | **Pulmonology Group** |
| Coralie Viollet | AstraZeneca, Cambridge, United Kingdom | Clinical Groups | **Pulmonology Group** |
| Susan Eaton | Biogen, Cambridge, MA, United States | Clinical Groups | **Pulmonology Group** |
| Hubert Chen | Genentech, San Francisco, CA, United States | Clinical Groups | **Pulmonology Group** |
| Rion Pendergrass | Genentech, San Francisco, CA, United States | Clinical Groups | **Pulmonology Group** |
| Natalie Bowers | Genentech, San Francisco, CA, United States | Clinical Groups | **Pulmonology Group** |
| Joanna Betts | GlaxoSmithKline, Brentford, United Kingdom | Clinical Groups | **Pulmonology Group** |

| | | | |
|---|---|---|---|
| Kirsi Auro | GlaxoSmithKline, Espoo, Finland | Clinical Groups | **Pulmonology Group** |
| Rajashree Mishra | GlaxoSmithKline, Brentford, United Kingdom | Clinical Groups | **Pulmonology Group** |
| Majd Mouded | Novartis, Basel, Switzerland | Clinical Groups | **Pulmonology Group** |
| Debby Ngo | Novartis, Basel, Switzerland | Clinical Groups | **Pulmonology Group** |
| Teemu Niiranen | Finnish Institute for Health and Welfare (THL), Helsinki, Finland | Clinical Groups | **Cardiometabolic Diseases Group** |
| Felix Vaura | Finnish Institute for Health and Welfare (THL), Helsinki, Finland | Clinical Groups | **Cardiometabolic Diseases Group** |
| Veikko Salomaa | Finnish Institute for Health and Welfare (THL), Helsinki, Finland | Clinical Groups | **Cardiometabolic Diseases Group** |
| Kaj Metsärinne | Hospital District of Southwest Finland, Turku, Finland | Clinical Groups | **Cardiometabolic Diseases Group** |
| Jenni Aittokallio | Hospital District of Southwest Finland, Turku, Finland | Clinical Groups | **Cardiometabolic Diseases Group** |
| Mika Kähönen | Pirkanmaa Hospital District, Tampere, Finland | Clinical Groups | **Cardiometabolic Diseases Group** |
| Jussi Hernesniemi | Pirkanmaa Hospital District, Tampere, Finland | Clinical Groups | **Cardiometabolic Diseases Group** |
| Daniel Gordin | Hospital District of Helsinki and Uusimaa, Helsinki, Finland | Clinical Groups | **Cardiometabolic Diseases Group** |
| Juha Sinisalo | Hospital District of Helsinki and Uusimaa, Helsinki, Finland | Clinical Groups | **Cardiometabolic Diseases Group** |
| Marja-Riitta Taskinen | Hospital District of Helsinki and Uusimaa, Helsinki, Finland | Clinical Groups | **Cardiometabolic Diseases Group** |
| Tiinamaija Tuomi | Hospital District of Helsinki and Uusimaa, Helsinki, Finland | Clinical Groups | **Cardiometabolic Diseases Group** |
| Timo Hiltunen | Hospital District of Helsinki and Uusimaa, Helsinki, Finland | Clinical Groups | **Cardiometabolic Diseases Group** |
| Jari Laukkanen | Central Finland Health Care District, Jyväskylä, Finland | Clinical Groups | **Cardiometabolic Diseases Group** |
| Amanda Elliott | Institute for Molecular Medicine Finland (FIMM), HiLIFE, University of Helsinki, Helsinki, Finland; Broad Institute, Cambridge, MA, USA and Massachusetts General Hospital, Boston, MA, USA | Clinical Groups | **Cardiometabolic Diseases Group** |
| Mary Pat Reeve | Institute for Molecular Medicine Finland (FIMM), HiLIFE, University of Helsinki, Helsinki, Finland | Clinical Groups | **Cardiometabolic Diseases Group** |
| Sanni Ruotsalainen | Institute for Molecular Medicine Finland (FIMM), HiLIFE, University of Helsinki, Helsinki, Finland | Clinical Groups | **Cardiometabolic Diseases Group** |
| Dirk Paul | Astra Zeneca, Cambridge, United Kingdom | Clinical Groups | **Cardiometabolic Diseases Group** |
| Natalie Bowers | Genentech, San Francisco, CA, United States | Clinical Groups | **Cardiometabolic Diseases Group** |
| Rion Pendergrass | Genentech, San Francisco, CA, United States | Clinical Groups | **Cardiometabolic Diseases Group** |
| Audrey Chu | GlaxoSmithKline, Brentford, United Kingdom | Clinical Groups | **Cardiometabolic Diseases Group** |
| Kirsi Auro | GlaxoSmithKline, Espoo, Finland | Clinical Groups | **Cardiometabolic Diseases Group** |
| Dermot Reilly | Janssen Research & Development, LLC, Boston, MA, United States | Clinical Groups | **Cardiometabolic Diseases Group** |
| Mike Mendelson | Novartis, Boston, MA, United States | Clinical Groups | **Cardiometabolic Diseases Group** |
| Jaakko Parkkinen | Pfizer, New York, NY, United States | Clinical Groups | **Cardiometabolic Diseases Group** |
| Melissa Miller | Pfizer, New York, NY, United States | Clinical Groups | **Cardiometabolic Diseases Group** |
| Tuomo Meretoja | Department of Breast Surgery, Helsinki University Hospital Comprehensive Cancer Center and University of Helsinki, Helsinki, Finland | Clinical Groups | **Oncology Group** |

| | | | |
|---|---|---|---|
| Heikki Joensuu | Department of Oncology, Helsinki University Hospital Comprehensive Cancer Center and University of Helsinki, Helsinki, Finland | Clinical Groups | **Oncology Group** |
| Olli Carpén | Hospital District of Helsinki and Uusimaa, Helsinki, Finland | Clinical Groups | **Oncology Group** |
| Johanna Mattson | Hospital District of Helsinki and Uusimaa, Helsinki, Finland | Clinical Groups | **Oncology Group** |
| Eveliina Salminen | Hospital District of Helsinki and Uusimaa, Helsinki, Finland | Clinical Groups | **Oncology Group** |
| Annika Auranen | Pirkanmaa Hospital District , Tampere, Finland | Clinical Groups | **Oncology Group** |
| Peeter Karihtala | Department of Oncology, Helsinki University Hospital Comprehensive Cancer Center and University of Helsinki, Helsinki, Finland | Clinical Groups | **Oncology Group** |
| Päivi Auvinen | Northern Savo Hospital District, Kuopio, Finland | Clinical Groups | **Oncology Group** |
| Klaus Elenius | Hospital District of Southwest Finland, Turku, Finland | Clinical Groups | **Oncology Group** |
| Johanna Schleutker | Hospital District of Southwest Finland, Turku, Finland | Clinical Groups | **Oncology Group** |
| Esa Pitkänen | Institute for Molecular Medicine Finland (FIMM), HiLIFE, University of Helsinki, Helsinki, Finland | Clinical Groups | **Oncology Group** |
| Nina Mars | Institute for Molecular Medicine Finland (FIMM), HiLIFE, University of Helsinki, Helsinki, Finland | Clinical Groups | **Oncology Group** |
| Mark Daly | Institute for Molecular Medicine Finland (FIMM), HiLIFE, University of Helsinki, Helsinki, Finland; Broad Institute of MIT and Harvard; Massachusetts General Hospital | Clinical Groups | **Oncology Group** |
| Relja Popovic | Abbvie, Chicago, IL, United States | Clinical Groups | **Oncology Group** |
| Jeffrey Waring | Abbvie, Chicago, IL, United States | Clinical Groups | **Oncology Group** |
| Bridget Riley-Gillis | Abbvie, Chicago, IL, United States | Clinical Groups | **Oncology Group** |
| Anne Lehtonen | Abbvie, Chicago, IL, United States | Clinical Groups | **Oncology Group** |
| Margarete Fabre | AstraZeneca, Cambridge, United Kingdom | Clinical Groups | **Oncology Group** |
| Jennifer Schutzman | Genentech, San Francisco, CA, United States | Clinical Groups | **Oncology Group** |
| Natalie Bowers | Genentech, San Francisco, CA, United States | Clinical Groups | **Oncology Group** |
| Rion Pendergrass | Genentech, San Francisco, CA, United States | Clinical Groups | **Oncology Group** |
| Diptee Kulkarni | GlaxoSmithKline, Brentford, United Kingdom | Clinical Groups | **Oncology Group** |
| Kirsi Auro | GlaxoSmithKline, Espoo, Finland | Clinical Groups | **Oncology Group** |
| Alessandro Porello | Janssen Research & Development, LLC, Spring House, PA, United States | Clinical Groups | **Oncology Group** |
| Andrey Loboda | Merck, Kenilworth, NJ, United States | Clinical Groups | **Oncology Group** |
| Heli Lehtonen | Pfizer, New York, NY, United States | Clinical Groups | **Oncology Group** |
| Stefan McDonough | Pfizer, New York, NY, United States | Clinical Groups | **Oncology Group** |
| Sauli Vuoti | Janssen-Cilag Oy, Espoo, Finland | Clinical Groups | **Oncology Group** |
| Kai Kaarniranta | Northern Savo Hospital District, Kuopio, Finland; Department of Molecular Genetics, University of Lodz, Lodz, Poland | Clinical Groups | **Opthalmology Group** |
| Joni A Turunen | Helsinki University Hospital and University of Helsinki, Helsinki, Finland; Eye Genetics Group, Folkhälsan Research Center, Helsinki, Finland | Clinical Groups | **Opthalmology Group** |
| Terhi Ollila | Hospital District of Helsinki and Uusimaa, Helsinki, Finland | Clinical Groups | **Opthalmology Group** |

| | | | |
|---|---|---|---|
| Hannu Uusitalo | Pirkanmaa Hospital District, Tampere, Finland | Clinical Groups | **Opthalmology Group** |
| Juha Karjalainen | Institute for Molecular Medicine Finland (FIMM), HiLIFE, University of Helsinki, Helsinki, Finland | Clinical Groups | **Opthalmology Group** |
| Esa Pitkänen | Institute for Molecular Medicine Finland (FIMM), HiLIFE, University of Helsinki, Helsinki, Finland | Clinical Groups | **Opthalmology Group** |
| Mengzhen Liu | Abbvie, Chicago, IL, United States | Clinical Groups | **Opthalmology Group** |
| Heiko Runz | Biogen, Cambridge, MA, United States | Clinical Groups | **Opthalmology Group** |
| Stephanie Loomis | Biogen, Cambridge, MA, United States | Clinical Groups | **Opthalmology Group** |
| Erich Strauss | Genentech, San Francisco, CA, United States | Clinical Groups | **Opthalmology Group** |
| Natalie Bowers | Genentech, San Francisco, CA, United States | Clinical Groups | **Opthalmology Group** |
| Hao Chen | Genentech, San Francisco, CA, United States | Clinical Groups | **Opthalmology Group** |
| Rion Pendergrass | Genentech, San Francisco, CA, United States | Clinical Groups | **Opthalmology Group** |
| Kaisa Tasanen | Northern Ostrobothnia Hospital District, Oulu, Finland | Clinical Groups | **Dermatology Group** |
| Laura Huilaja | Northern Ostrobothnia Hospital District, Oulu, Finland | Clinical Groups | **Dermatology Group** |
| Katariina Hannula-Jouppi | Hospital District of Helsinki and Uusimaa, Helsinki, Finland | Clinical Groups | **Dermatology Group** |
| Teea Salmi | Pirkanmaa Hospital District, Tampere, Finland | Clinical Groups | **Dermatology Group** |
| Sirkku Peltonen | Hospital District of Southwest Finland, Turku, Finland | Clinical Groups | **Dermatology Group** |
| Leena Koulu | Hospital District of Southwest Finland, Turku, Finland | Clinical Groups | **Dermatology Group** |
| Nizar Smaoui | Abbvie, Chicago, IL, United States | Clinical Groups | **Dermatology Group** |
| Fedik Rahimov | Abbvie, Chicago, IL, United States | Clinical Groups | **Dermatology Group** |
| Anne Lehtonen | Abbvie, Chicago, IL, United States | Clinical Groups | **Dermatology Group** |
| David Choy | Genentech, San Francisco, CA, United States | Clinical Groups | **Dermatology Group** |
| Rion Pendergrass | Genentech, San Francisco, CA, United States | Clinical Groups | **Dermatology Group** |
| Dawn Waterworth | Janssen Research & Development, LLC, Spring House, PA, United States | Clinical Groups | **Dermatology Group** |
| Kirsi Kalpala | Pfizer, New York, NY, United States | Clinical Groups | **Dermatology Group** |
| Ying Wu | Pfizer, New York, NY, United States | Clinical Groups | **Dermatology Group** |
| Pirkko Pussinen | Hospital District of Helsinki and Uusimaa, Helsinki, Finland | Clinical Groups | **Odontology Group** |
| Aino Salminen | Hospital District of Helsinki and Uusimaa, Helsinki, Finland | Clinical Groups | **Odontology Group** |
| Tuula Salo | Hospital District of Helsinki and Uusimaa, Helsinki, Finland | Clinical Groups | **Odontology Group** |
| David Rice | Hospital District of Helsinki and Uusimaa, Helsinki, Finland | Clinical Groups | **Odontology Group** |
| Pekka Nieminen | Hospital District of Helsinki and Uusimaa, Helsinki, Finland | Clinical Groups | **Odontology Group** |
| Ulla Palotie | Hospital District of Helsinki and Uusimaa, Helsinki, Finland | Clinical Groups | **Odontology Group** |
| Maria Siponen | Northern Savo Hospital District, Kuopio, Finland | Clinical Groups | **Odontology Group** |

| | | | |
|---|---|---|---|
| Liisa Suominen | Northern Savo Hospital District, Kuopio, Finland | Clinical Groups | **Odontology Group** |
| Päivi Mäntylä | Northern Savo Hospital District, Kuopio, Finland | Clinical Groups | **Odontology Group** |
| Ulvi Gursoy | Hospital District of Southwest Finland, Turku, Finland | Clinical Groups | **Odontology Group** |
| Vuokko Anttonen | Northern Ostrobothnia Hospital District, Oulu, Finland | Clinical Groups | **Odontology Group** |
| Kirsi Sipilä | Research Unit of Oral Health Sciences Faculty of Medicine, University of Oulu, Oulu, Finland; Medical Research Center, Oulu, Oulu University Hospital and University of Oulu, Oulu, Finland | Clinical Groups | **Odontology Group** |
| Rion Pendergrass | Genentech, San Francisco, CA, United States | Clinical Groups | **Odontology Group** |
| Hannele Laivuori | Institute for Molecular Medicine Finland (FIMM), HiLIFE, University of Helsinki, Helsinki, Finland | Clinical Groups | **Women's Health and Reproduction Group** |
| Venla Kurra | Pirkanmaa Hospital District, Tampere, Finland | Clinical Groups | **Women's Health and Reproduction Group** |
| Laura Kotaniemi-Talonen | Pirkanmaa Hospital District, Tampere, Finland | Clinical Groups | **Women's Health and Reproduction Group** |
| Oskari Heikinheimo | Hospital District of Helsinki and Uusimaa, Helsinki, Finland | Clinical Groups | **Women's Health and Reproduction Group** |
| Ilkka Kalliala | Hospital District of Helsinki and Uusimaa, Helsinki, Finland | Clinical Groups | **Women's Health and Reproduction Group** |
| Lauri Aaltonen | Hospital District of Helsinki and Uusimaa, Helsinki, Finland | Clinical Groups | **Women's Health and Reproduction Group** |
| Varpu Jokimaa | Hospital District of Southwest Finland, Turku, Finland | Clinical Groups | **Women's Health and Reproduction Group** |
| Johannes Kettunen | Northern Ostrobothnia Hospital District, Oulu, Finland | Clinical Groups | **Women's Health and Reproduction Group** |
| Marja Vääräsmäki | Northern Ostrobothnia Hospital District, Oulu, Finland | Clinical Groups | **Women's Health and Reproduction Group** |
| Outi Uimari | Northern Ostrobothnia Hospital District, Oulu, Finland | Clinical Groups | **Women's Health and Reproduction Group** |
| Laure Morin-Papunen | Northern Ostrobothnia Hospital District, Oulu, Finland | Clinical Groups | **Women's Health and Reproduction Group** |
| Maarit Niinimäki | Northern Ostrobothnia Hospital District, Oulu, Finland | Clinical Groups | **Women's Health and Reproduction Group** |
| Terhi Piltonen | Northern Ostrobothnia Hospital District, Oulu, Finland | Clinical Groups | **Women's Health and Reproduction** |

| | | | |
|---|---|---|---|
| | | | Group |
| Katja Kivinen | Institute for Molecular Medicine Finland (FIMM), HiLIFE, University of Helsinki, Helsinki, Finland | Clinical Groups | **Women's Health and Reproduction Group** |
| Elisabeth Widen | Institute for Molecular Medicine Finland (FIMM), HiLIFE, University of Helsinki, Helsinki, Finland | Clinical Groups | **Women's Health and Reproduction Group** |
| Taru Tukiainen | Institute for Molecular Medicine Finland (FIMM), HiLIFE, University of Helsinki, Helsinki, Finland | Clinical Groups | **Women's Health and Reproduction Group** |
| Mary Pat Reeve | Institute for Molecular Medicine Finland (FIMM), HiLIFE, University of Helsinki, Helsinki, Finland | Clinical Groups | **Women's Health and Reproduction Group** |
| Mark Daly | Institute for Molecular Medicine Finland (FIMM), HiLIFE, University of Helsinki, Helsinki, Finland; Broad Institute of MIT and Harvard; Massachusetts General Hospital | Clinical Groups | **Women's Health and Reproduction Group** |
| Niko Välimäki | University of Helsinki, Helsinki, Finland | Clinical Groups | **Women's Health and Reproduction Group** |
| Eija Laakkonen | University of Jyväskylä, Jyväskylä, Finland | Clinical Groups | **Women's Health and Reproduction Group** |
| Jaakko Tyrmi | University of Oulu, Oulu, Finland / University of Tampere, Tampere, Finland | Clinical Groups | **Women's Health and Reproduction Group** |
| Heidi Silven | University of Oulu, Oulu, Finland | Clinical Groups | **Women's Health and Reproduction Group** |
| Eeva Sliz | University of Oulu, Oulu, Finland | Clinical Groups | **Women's Health and Reproduction Group** |
| Riikka Arffman | University of Oulu, Oulu, Finland | Clinical Groups | **Women's Health and Reproduction Group** |
| Susanna Savukoski | University of Oulu, Oulu, Finland | Clinical Groups | **Women's Health and Reproduction Group** |
| Triin Laisk | Estonian biobank, Tartu, Estonia | Clinical Groups | **Women's Health and Reproduction Group** |
| Natalia Pujol | Estonian biobank, Tartu, Estonia | Clinical Groups | **Women's Health and Reproduction Group** |
| Mengzhen Liu | Abbvie, Chicago, IL, United States | Clinical Groups | **Women's Health and Reproduction Group** |
| Bridget Riley-Gillis | Abbvie, Chicago, IL, United States | Clinical Groups | **Women's Health and Reproduction** |

| | | | |
|---|---|---|---|
| | | | **Group** |
| Rion Pendergrass | Genentech, San Francisco, CA, United States | Clinical Groups | **Women's Health and Reproduction Group** |
| Janet Kumar | GlaxoSmithKline, Collegeville, PA, United States | Clinical Groups | **Women's Health and Reproduction Group** |
| Kirsi Auro | GlaxoSmithKline, Espoo, Finland | Clinical Groups | **Women's Health and Reproduction Group** |
| Iiris Hovatta | University of Helsinki, Finland | Clinical Groups | **Depression group** |
| Chia-Yen Chen | Biogen, Cambridge, MA, United States | Clinical Groups | **Depression group** |
| Erkki Isometsä | Hospital District of Helsinki and Uusimaa, Helsinki, Finland | Clinical Groups | **Depression group** |
| Hanna Ollila | Institute for Molecular Medicine Finland (FIMM), HiLIFE, University of Helsinki, Helsinki, Finland | Clinical Groups | **Depression group** |
| Jaana Suvisaari | Finnish Institute for Health and Welfare (THL), Helsinki, Finland | Clinical Groups | **Depression group** |
| Antti Mäkitie | Department of Otorhinolaryngology - Head and Neck Surgery, University of Helsinki and Helsinki University Hospital, Helsinki, Finland | Clinical Groups | **ENT (ear, nose and throath) Group** |
| Argyro Bizaki-Vallaskangas | Pirkanmaa Hospital District, Tampere, Finland | Clinical Groups | **ENT (ear, nose and throath) Group** |
| Sanna Toppila-Salmi | University of Eastern Finland and Kuopio University Hospital, Department of Otorhinolaryngology, Kuopio, Finland and Department of Allergy, Helsinki University Hospital and University of Helsinki, Finland | Clinical Groups | **ENT (ear, nose and throath) Group** |
| Tytti Willberg | Hospital District of Southwest Finland, Turku, Finland | Clinical Groups | **ENT (ear, nose and throath) Group** |
| Elmo Saarentaus | Institute for Molecular Medicine Finland (FIMM), HiLIFE, University of Helsinki, Helsinki, Finland | Clinical Groups | **ENT (ear, nose and throath) Group** |
| Antti Aarnisalo | Hospital District of Helsinki and Uusimaa, Helsinki, Finland | Clinical Groups | **ENT (ear, nose and throath) Group** |
| Eveliina Salminen | Hospital District of Helsinki and Uusimaa, Helsinki, Finland | Clinical Groups | **ENT (ear, nose and throath) Group** |
| Elisa Rahikkala | Northern Ostrobothnia Hospital District, Oulu, Finland | Clinical Groups | **ENT (ear, nose and throath) Group** |
| Johannes Kettunen | Northern Ostrobothnia Hospital District, Oulu, Finland | Clinical Groups | **ENT (ear, nose and throath) Group** |
| Kristiina Aittomäki | Department of Medical Genetics, Helsinki University Central Hospital, Helsinki, Finland | Clinical Groups | **POI (premature ovarian failure) Group** |
| Fredrik Åberg | Transplantation and Liver Surgery Clinic, Helsinki University Hospital, Helsinki University, Helsinki, Finland | Clinical Groups | **LiverScore Group** |
| Mitja Kurki | Institute for Molecular Medicine Finland (FIMM), HiLIFE, University of Helsinki, Helsinki, Finland; Broad Institute, Cambridge, MA, United States | FinnGen Analysis working group | FinnGen Analysis working group |
| Samuli Ripatti | Institute for Molecular Medicine Finland (FIMM), HiLIFE, University of Helsinki, Helsinki, Finland | FinnGen Analysis working group | FinnGen Analysis working group |

| | | | |
|---|---|---|---|
| Mark Daly | Institute for Molecular Medicine, Finland (FIMM), HiLIFE, University of Helsinki, Helsinki, Finland; Broad Institute of MIT and Harvard; Massachusetts General Hospital | FinnGen Analysis working group | FinnGen Analysis working group |
| Juha Karjalainen | Institute for Molecular Medicine Finland (FIMM), HiLIFE, University of Helsinki, Helsinki, Finland | FinnGen Analysis working group | FinnGen Analysis working group |
| Aki Havulinna | Institute for Molecular Medicine Finland (FIMM), HiLIFE, University of Helsinki, Helsinki, Finland; Finnish Institute for Health and Welfare (THL), Helsinki, Finland | FinnGen Analysis working group | FinnGen Analysis working group |
| Juha Mehtonen | Institute for Molecular Medicine Finland (FIMM), HiLIFE, University of Helsinki, Helsinki, Finland | FinnGen Analysis working group | FinnGen Analysis working group |
| Priit Palta | Institute for Molecular Medicine Finland (FIMM), HiLIFE, University of Helsinki, Helsinki, Finland | FinnGen Analysis working group | FinnGen Analysis working group |
| Shabbeer Hassan | Institute for Molecular Medicine Finland (FIMM), HiLIFE, University of Helsinki, Helsinki, Finland | FinnGen Analysis working group | FinnGen Analysis working group |
| Pietro Della Briotta Parolo | Institute for Molecular Medicine Finland (FIMM), HiLIFE, University of Helsinki, Helsinki, Finland | FinnGen Analysis working group | FinnGen Analysis working group |
| Wei Zhou | Broad Institute, Cambridge, MA, United States | FinnGen Analysis working group | FinnGen Analysis working group |
| Mutaamba Maasha | Broad Institute, Cambridge, MA, United States | FinnGen Analysis working group | FinnGen Analysis working group |
| Shabbeer Hassan | Institute for Molecular Medicine Finland (FIMM), HiLIFE, University of Helsinki, Helsinki, Finland | FinnGen Analysis working group | FinnGen Analysis working group |
| Susanna Lemmelä | Institute for Molecular Medicine Finland (FIMM), HiLIFE, University of Helsinki, Helsinki, Finland | FinnGen Analysis working group | FinnGen Analysis working group |
| Manuel Rivas | University of Stanford, Stanford, CA, United States | FinnGen Analysis working group | FinnGen Analysis working group |
| Aarno Palotie | Institute for Molecular Medicine Finland (FIMM), HiLIFE, University of Helsinki, Helsinki, Finland | FinnGen Analysis working group | FinnGen Analysis working group |
| Aoxing Liu | Institute for Molecular Medicine Finland (FIMM), HiLIFE, University of Helsinki, Helsinki, Finland | FinnGen Analysis working group | FinnGen Analysis working group |
| Arto Lehisto | Institute for Molecular Medicine Finland (FIMM), HiLIFE, University of Helsinki, Helsinki, Finland | FinnGen Analysis working group | FinnGen Analysis working group |
| Andrea Ganna | Institute for Molecular Medicine Finland (FIMM), HiLIFE, University of Helsinki, Helsinki, Finland | FinnGen Analysis working group | FinnGen Analysis working group |

| | | | |
|---|---|---|---|
| Vincent Llorens | Institute for Molecular Medicine Finland (FIMM), HiLIFE, University of Helsinki, Helsinki, Finland | FinnGen Analysis working group | FinnGen Analysis working group |
| Hannele Laivuori | Institute for Molecular Medicine Finland (FIMM), HiLIFE, University of Helsinki, Helsinki, Finland | FinnGen Analysis working group | FinnGen Analysis working group |
| Taru Tukiainen | Institute for Molecular Medicine Finland (FIMM), HiLIFE, University of Helsinki, Helsinki, Finland | FinnGen Analysis working group | FinnGen Analysis working group |
| Mary Pat Reeve | Institute for Molecular Medicine Finland (FIMM), HiLIFE, University of Helsinki, Helsinki, Finland | FinnGen Analysis working group | FinnGen Analysis working group |
| Henrike Heyne | Institute for Molecular Medicine Finland (FIMM), HiLIFE, University of Helsinki, Helsinki, Finland | FinnGen Analysis working group | FinnGen Analysis working group |
| Nina Mars | Institute for Molecular Medicine Finland (FIMM), HiLIFE, University of Helsinki, Helsinki, Finland | FinnGen Analysis working group | FinnGen Analysis working group |
| Joel Rämö | Institute for Molecular Medicine Finland (FIMM), HiLIFE, University of Helsinki, Helsinki, Finland | FinnGen Analysis working group | FinnGen Analysis working group |
| Elmo Saarentaus | Institute for Molecular Medicine Finland (FIMM), HiLIFE, University of Helsinki, Helsinki, Finland | FinnGen Analysis working group | FinnGen Analysis working group |
| Hanna Ollila | Institute for Molecular Medicine Finland (FIMM), HiLIFE, University of Helsinki, Helsinki, Finland | FinnGen Analysis working group | FinnGen Analysis working group |
| Rodos Rodosthenous | Institute for Molecular Medicine Finland (FIMM), HiLIFE, University of Helsinki, Helsinki, Finland | FinnGen Analysis working group | FinnGen Analysis working group |
| Satu Strausz | Institute for Molecular Medicine Finland (FIMM), HiLIFE, University of Helsinki, Helsinki, Finland | FinnGen Analysis working group | FinnGen Analysis working group |
| Tuula Palotie | University of Helsinki and Hospital District of Helsinki and Uusimaa, Helsinki, Finland | FinnGen Analysis working group | FinnGen Analysis working group |
| Kimmo Palin | University of Helsinki, Helsinki, Finland | FinnGen Analysis working group | FinnGen Analysis working group |
| Javier Garcia-Tabuenca | University of Tampere, Tampere, Finland | FinnGen Analysis working group | FinnGen Analysis working group |
| Harri Siirtola | University of Tampere, Tampere, Finland | FinnGen Analysis working group | FinnGen Analysis working group |
| Tuomo Kiiskinen | Institute for Molecular Medicine Finland (FIMM), HiLIFE, University of Helsinki, Helsinki, Finland | FinnGen Analysis working group | FinnGen Analysis working group |

| | | | |
|---|---|---|---|
| Jiwoo Lee | Institute for Molecular Medicine Finland (FIMM), HiLIFE, University of Helsinki, Helsinki, Finland; Broad Institute, Cambridge, MA, United States | **FinnGen Analysis working group** | **FinnGen Analysis working group** |
| Kristin Tsuo | Institute for Molecular Medicine Finland (FIMM), HiLIFE, University of Helsinki, Helsinki, Finland; Broad Institute, Cambridge, MA, United States | **FinnGen Analysis working group** | **FinnGen Analysis working group** |
| Amanda Elliott | Institute for Molecular Medicine Finland (FIMM), HiLIFE, University of Helsinki, Helsinki, Finland; Broad Institute, Cambridge, MA, USA and Massachusetts General Hospital, Boston, MA, USA | **FinnGen Analysis working group** | **FinnGen Analysis working group** |
| Kati Kristiansson | THL Biobank / Finnish Institute for Health and Welfare (THL), Helsinki, Finland | **FinnGen Analysis working group** | **FinnGen Analysis working group** |
| Mikko Arvas | Finnish Red Cross Blood Service / Finnish Hematology Registry and Clinical Biobank, Helsinki, Finland | **FinnGen Analysis working group** | **FinnGen Analysis working group** |
| Kati Hyvärinen | Finnish Red Cross Blood Service, Helsinki, Finland | **FinnGen Analysis working group** | **FinnGen Analysis working group** |
| Jarmo Ritari | Finnish Red Cross Blood Service, Helsinki, Finland | **FinnGen Analysis working group** | **FinnGen Analysis working group** |
| Olli Carpén | Helsinki Biobank / Helsinki University and Hospital District of Helsinki and Uusimaa, Helsinki | **FinnGen Analysis working group** | **FinnGen Analysis working group** |
| Johannes Kettunen | Northern Finland Biobank Borealis / University of Oulu / Northern Ostrobothnia Hospital District, Oulu, Finland | **FinnGen Analysis working group** | **FinnGen Analysis working group** |
| Katri Pylkäs | University of Oulu, Oulu, Finland | **FinnGen Analysis working group** | **FinnGen Analysis working group** |
| Eeva Sliz | University of Oulu, Oulu, Finland | **FinnGen Analysis working group** | **FinnGen Analysis working group** |
| Minna Karjalainen | University of Oulu, Oulu, Finland | **FinnGen Analysis working group** | **FinnGen Analysis working group** |
| Tuomo Mantere | Northern Finland Biobank Borealis / University of Oulu / Northern Ostrobothnia Hospital District, Oulu, Finland | **FinnGen Analysis working group** | **FinnGen Analysis working group** |
| Eeva Kangasniemi | Finnish Clinical Biobank Tampere / University of Tampere / Pirkanmaa Hospital District, Tampere, Finland | **FinnGen Analysis working group** | **FinnGen Analysis working group** |
| Sami Heikkinen | University of Eastern Finland, Kuopio, Finland | **FinnGen Analysis working group** | **FinnGen Analysis working group** |
| Arto Mannermaa | Biobank of Eastern Finland / University of Eastern Finland / Northern Savo Hospital District, Kuopio, Finland | **FinnGen Analysis working group** | **FinnGen Analysis working group** |

| | | | |
|---|---|---|---|
| Eija Laakkonen | University of Jyväskylä, Jyväskylä, Finland | FinnGen Analysis working group | FinnGen Analysis working group |
| Nina Pitkänen | Auria Biobank / University of Turku / Hospital District of Southwest Finland, Turku, Finland | FinnGen Analysis working group | FinnGen Analysis working group |
| Samuel Lessard | Translational Sciences, Sanofi R&D, Framingham, MA, USA | FinnGen Analysis working group | FinnGen Analysis working group |
| Clément Chatelain | Translational Sciences, Sanofi R&D, Framingham, MA, USA | FinnGen Analysis working group | FinnGen Analysis working group |
| Lila Kallio | Auria Biobank / University of Turku / Hospital District of Southwest Finland, Turku, Finland | Biobank directors | Biobank directors |
| Tiina Wahlfors | THL Biobank / Finnish Institute for Health and Welfare (THL), Helsinki, Finland | Biobank directors | Biobank directors |
| Jukka Partanen | Finnish Red Cross Blood Service / Finnish Hematology Registry and Clinical Biobank, Helsinki, Finland | Biobank directors | Biobank directors |
| Eero Punkka | Helsinki Biobank / Helsinki University and Hospital District of Helsinki and Uusimaa, Helsinki | Biobank directors | Biobank directors |
| Raisa Serpi | Northern Finland Biobank Borealis / University of Oulu / Northern Ostrobothnia Hospital District, Oulu, Finland | Biobank directors | Biobank directors |
| Sanna Siltanen | Finnish Clinical Biobank Tampere / University of Tampere / Pirkanmaa Hospital District, Tampere, Finland | Biobank directors | Biobank directors |
| Veli-Matti Kosma | Biobank of Eastern Finland / University of Eastern Finland / Northern Savo Hospital District, Kuopio, Finland | Biobank directors | Biobank directors |
| Teijo Kuopio | Central Finland Biobank / University of Jyväskylä / Central Finland Health Care District, Jyväskylä, Finland | Biobank directors | Biobank directors |
| Anu Jalanko | Institute for Molecular Medicine Finland (FIMM), HiLIFE, University of Helsinki, Helsinki, Finland | FinnGen Teams | **Administration** |
| Huei-Yi Shen | Institute for Molecular Medicine Finland (FIMM), HiLIFE, University of Helsinki, Helsinki, Finland | FinnGen Teams | **Administration** |
| Risto Kajanne | Institute for Molecular Medicine Finland (FIMM), HiLIFE, University of Helsinki, Helsinki, Finland | FinnGen Teams | **Administration** |
| Mervi Aavikko | Institute for Molecular Medicine Finland (FIMM), HiLIFE, University of Helsinki, Helsinki, Finland | FinnGen Teams | **Administration** |
| Helen Cooper | Institute for Molecular Medicine Finland (FIMM), HiLIFE, University of Helsinki, Helsinki, Finland | FinnGen Teams | **Administration** |
| Denise Öller | Institute for Molecular Medicine Finland (FIMM), HiLIFE, University of Helsinki, Helsinki, Finland | FinnGen Teams | **Administration** |
| Rasko Leinonen | Institute for Molecular Medicine Finland (FIMM), HiLIFE, University of Helsinki, Helsinki, Finland; European Molecular Biology Laboratory, European Bioinformatics Institute, Cambridge, UK | FinnGen Teams | **Administration** |
| Henna Palin | Finnish Clinical Biobank Tampere / University of Tampere / Pirkanmaa Hospital District, Tampere, Finland | FinnGen Teams | **Administration** |
| Malla-Maria Linna | Helsinki Biobank / Helsinki University and Hospital District of Helsinki and Uusimaa, Helsinki | FinnGen Teams | **Administration** |
| Mitja Kurki | Institute for Molecular Medicine Finland (FIMM), HiLIFE, University of Helsinki, Helsinki, Finland; Broad Institute, Cambridge, MA, United States | FinnGen Teams | **Analysis** |
| Juha Karjalainen | Institute for Molecular Medicine Finland (FIMM), HiLIFE, University of Helsinki, Helsinki, Finland | FinnGen Teams | **Analysis** |
| Pietro Della Briotta Parolo | Institute for Molecular Medicine Finland (FIMM), HiLIFE, University of Helsinki, Helsinki, Finland | FinnGen Teams | **Analysis** |
| Arto Lehisto | Institute for Molecular Medicine Finland (FIMM), HiLIFE, University of Helsinki, Helsinki, Finland | FinnGen Teams | **Analysis** |
| Juha Mehtonen | Institute for Molecular Medicine Finland (FIMM), HiLIFE, University of Helsinki, Helsinki, Finland | FinnGen Teams | **Analysis** |
| Wei Zhou | Broad Institute, Cambridge, MA, United States | FinnGen Teams | **Analysis** |

| | | | |
|---|---|---|---|
| Masahiro Kanai | Broad Institute, Cambridge, MA, United States | FinnGen Teams | **Analysis** |
| Mutaamba Maasha | Broad Institute, Cambridge, MA, United States | FinnGen Teams | **Analysis** |
| Zhili Zheng | Broad Institute, Cambridge, MA, United States | FinnGen Teams | **Analysis** |
| Hannele Laivuori | Institute for Molecular Medicine Finland (FIMM), HiLIFE, University of Helsinki, Helsinki, Finland | FinnGen Teams | **Clinical Endpoint Development** |
| Aki Havulinna | Institute for Molecular Medicine Finland (FIMM), HiLIFE, University of Helsinki, Helsinki, Finland; Finnish Institute for Health and Welfare (THL), Helsinki, Finland | FinnGen Teams | **Clinical Endpoint Development** |
| Susanna Lemmelä | Institute for Molecular Medicine Finland (FIMM), HiLIFE, University of Helsinki, Helsinki, Finland | FinnGen Teams | **Clinical Endpoint Development** |
| Tuomo Kiiskinen | Institute for Molecular Medicine Finland (FIMM), HiLIFE, University of Helsinki, Helsinki, Finland | FinnGen Teams | **Clinical Endpoint Development** |
| L. Elisa Lahtela | Institute for Molecular Medicine Finland (FIMM), HiLIFE, University of Helsinki, Helsinki, Finland | FinnGen Teams | **Clinical Endpoint Development** |
| Mari Kaunisto | Institute for Molecular Medicine Finland (FIMM), HiLIFE, University of Helsinki, Helsinki, Finland | FinnGen Teams | **Communication** |
| Elina Kilpeläinen | Institute for Molecular Medicine Finland (FIMM), HiLIFE, University of Helsinki, Helsinki, Finland | FinnGen Teams | **E-Science** |
| Timo P. Sipilä | Institute for Molecular Medicine Finland (FIMM), HiLIFE, University of Helsinki, Helsinki, Finland | FinnGen Teams | **E-Science** |
| Oluwaseun Alexander Dada | Institute for Molecular Medicine Finland (FIMM), HiLIFE, University of Helsinki, Helsinki, Finland | FinnGen Teams | **E-Science** |
| Awaisa Ghazal | Institute for Molecular Medicine Finland (FIMM), HiLIFE, University of Helsinki, Helsinki, Finland | FinnGen Teams | **E-Science** |
| Anastasia Kytölä | Institute for Molecular Medicine Finland (FIMM), HiLIFE, University of Helsinki, Helsinki, Finland | FinnGen Teams | **E-Science** |
| Rigbe Weldatsadik | Institute for Molecular Medicine Finland (FIMM), HiLIFE, University of Helsinki, Helsinki, Finland | FinnGen Teams | **E-Science** |
| Sanni Ruotsalainen | Institute for Molecular Medicine Finland (FIMM), HiLIFE, University of Helsinki, Helsinki, Finland | FinnGen Teams | **E-Science** |
| Kati Donner | Institute for Molecular Medicine Finland (FIMM), HiLIFE, University of Helsinki, Helsinki, Finland | FinnGen Teams | **Genotyping** |
| Timo P. Sipilä | Institute for Molecular Medicine Finland (FIMM), HiLIFE, University of Helsinki, Helsinki, Finland | FinnGen Teams | **Genotyping** |
| Anu Loukola | Helsinki Biobank / Helsinki University and Hospital District of Helsinki and Uusimaa, Helsinki | FinnGen Teams | **Sample Collection Coordination** |
| Päivi Laiho | THL Biobank / Finnish Institute for Health and Welfare (THL), Helsinki, Finland | FinnGen Teams | **Sample Logistics** |
| Tuuli Sistonen | THL Biobank / Finnish Institute for Health and Welfare (THL), Helsinki, Finland | FinnGen Teams | **Sample Logistics** |
| Essi Kaiharju | THL Biobank / Finnish Institute for Health and Welfare (THL), Helsinki, Finland | FinnGen Teams | **Sample Logistics** |
| Markku Laukkanen | THL Biobank / Finnish Institute for Health and Welfare (THL), Helsinki, Finland | FinnGen Teams | **Sample Logistics** |
| Elina Järvensivu | THL Biobank / Finnish Institute for Health and Welfare (THL), Helsinki, Finland | FinnGen Teams | **Sample Logistics** |
| Sini Lähteenmäki | THL Biobank / Finnish Institute for Health and Welfare (THL), Helsinki, Finland | FinnGen Teams | **Sample Logistics** |
| Lotta Männikkö | THL Biobank / Finnish Institute for Health and Welfare (THL), Helsinki, Finland | FinnGen Teams | **Sample Logistics** |
| Regis Wong | THL Biobank / Finnish Institute for Health and Welfare (THL), Helsinki, Finland | FinnGen Teams | **Sample Logistics** |
| Auli Toivola | THL Biobank / Finnish Institute for Health and Welfare (THL), Helsinki, Finland | FinnGen Teams | **Sample Logistics** |
| Minna Brunfeldt | THL Biobank / Finnish Institute for Health and Welfare (THL), Helsinki, Finland | FinnGen Teams | **Registry Data Operations** |
| Hannele Mattsson | THL Biobank / Finnish Institute for Health and Welfare (THL), Helsinki, Finland | FinnGen Teams | **Registry Data Operations** |

| | | | |
|---|---|---|---|
| Kati Kristiansson | THL Biobank / Finnish Institute for Health and Welfare (THL), Helsinki, Finland | FinnGen Teams | **Registry Data Operations** |
| Susanna Lemmelä | Institute for Molecular Medicine Finland (FIMM), HiLIFE, University of Helsinki, Helsinki, Finland | FinnGen Teams | **Registry Data Operations** |
| Sami Koskelainen | THL Biobank / Finnish Institute for Health and Welfare (THL), Helsinki, Finland | FinnGen Teams | **Registry Data Operations** |
| Tero Hiekkalinna | THL Biobank / Finnish Institute for Health and Welfare (THL), Helsinki, Finland | FinnGen Teams | **Registry Data Operations** |
| Teemu Paajanen | THL Biobank / Finnish Institute for Health and Welfare (THL), Helsinki, Finland | FinnGen Teams | **Registry Data Operations** |
| Priit Palta | Institute for Molecular Medicine Finland (FIMM), HiLIFE, University of Helsinki, Helsinki, Finland | FinnGen Teams | **Sequencing Informatics** |
| Shuang Luo | Institute for Molecular Medicine Finland (FIMM), HiLIFE, University of Helsinki, Helsinki, Finland | FinnGen Teams | **Sequencing Informatics** |
| Tarja Laitinen | Pirkanmaa Hospital District, Tampere, Finland | FinnGen Teams | **Trajectory** |
| Mary Pat Reeve | Institute for Molecular Medicine Finland (FIMM), HiLIFE, University of Helsinki, Helsinki, Finland | FinnGen Teams | **Trajectory** |
| Shanmukha Sampath Padmanabhuni | Institute for Molecular Medicine Finland (FIMM), HiLIFE, University of Helsinki, Helsinki, Finland | FinnGen Teams | **Trajectory** |
| Marianna Niemi | University of Tampere, Tampere, Finland | FinnGen Teams | **Trajectory** |
| Harri Siirtola | University of Tampere, Tampere, Finland | FinnGen Teams | **Trajectory** |
| Javier Gracia-Tabuenca | University of Tampere, Tampere, Finland | FinnGen Teams | **Trajectory** |
| Mika Helminen | University of Tampere, Tampere, Finland | FinnGen Teams | **Trajectory** |
| Tiina Luukkaala | University of Tampere, Tampere, Finland | FinnGen Teams | **Trajectory** |
| Iida Vähätalo | University of Tampere, Tampere, Finland | FinnGen Teams | **Trajectory** |
| Jyrki Tammerluoto | Institute for Molecular Medicine Finland (FIMM), HiLIFE, University of Helsinki, Helsinki, Finland | FinnGen Teams | **Data protection officer** |
| Marco Hautalahti | Finnish Biobank Cooperative - FINBB | FinnGen Teams | **FINBB - Finnish biobank cooperative** |
| Johanna Mäkelä | Finnish Biobank Cooperative - FINBB | FinnGen Teams | **FINBB - Finnish biobank cooperative** |
| Sarah Smith | Finnish Biobank Cooperative - FINBB | FinnGen Teams | **FINBB - Finnish biobank cooperative** |
| Tom Southerington | Finnish Biobank Cooperative - FINBB | FinnGen Teams | **FINBB - Finnish biobank cooperative** |
| Petri Lehto | Finnish Biobank Cooperative - FINBB | FinnGen Teams | **FINBB - Finnish biobank cooperative** |

### Million Veteran Program Full Acknowledgement

**MVP Program Office**

- Sumitra Muralidhar, Ph.D., Program Director
US Department of Veterans Affairs, 810 Vermont Avenue NW, Washington, DC 20420
- Jennifer Moser, Ph.D., Associate Director, Scientific Programs
US Department of Veterans Affairs, 810 Vermont Avenue NW, Washington, DC 20420
- Jennifer E. Deen, B.S., Associate Director, Cohort & Public Relations
US Department of Veterans Affairs, 810 Vermont Avenue NW, Washington, DC 20420

**MVP Executive Committee**

- Co-Chair: Philip S. Tsao, Ph.D.
VA Palo Alto Health Care System, 3801 Miranda Avenue, Palo Alto, CA 94304
- Co-Chair: Sumitra Muralidhar, Ph.D.
US Department of Veterans Affairs, 810 Vermont Avenue NW, Washington, DC 20420
- J. Michael Gaziano, M.D., M.P.H.
VA Boston Healthcare System, 150 S. Huntington Avenue, Boston, MA 02130
- Elizabeth Hauser, Ph.D.
Durham VA Medical Center, 508 Fulton Street, Durham, NC 27705
- Amy Kilbourne, Ph.D., M.P.H.
VA HSR&D, 2215 Fuller Road, Ann Arbor, MI 48105
- Michael Matheny, M.D., M.S., M.P.H.
VA Tennessee Valley Healthcare System, 1310 24th Ave. South, Nashville, TN 37212
- Dave Oslin, M.D.
Philadelphia VA Medical Center, 3900 Woodland Avenue, Philadelphia, PA 19104

**MVP Co-Principal Investigators**

- J. Michael Gaziano, M.D., M.P.H.
VA Boston Healthcare System, 150 S. Huntington Avenue, Boston, MA 02130
- Philip S. Tsao, Ph.D.
VA Palo Alto Health Care System, 3801 Miranda Avenue, Palo Alto, CA 94304

**MVP Core Operations**

- Jessica V. Brewer, M.P.H., Director, MVP Recruitment & Enrollment
VA Boston Healthcare System, 150 S. Huntington Avenue, Boston, MA 02130
- Mary T. Brophy M.D., M.P.H., Director, VA Central Biorepository
VA Boston Healthcare System, 150 S. Huntington Avenue, Boston, MA 02130
- Kelly Cho, M.P.H, Ph.D., Director, MVP Phenomics Data Core
VA Boston Healthcare System, 150 S. Huntington Avenue, Boston, MA 02130

- Lori Churby, B.S., Director, MVP Regulatory Affairs
VA Palo Alto Health Care System, 3801 Miranda Avenue, Palo Alto, CA 94304
- Scott L. DuVall, Ph.D., Director, VA Informatics and Computing Infrastructure (VINCI)
VA Salt Lake City Health Care System, 500 Foothill Drive, Salt Lake City, UT 84148
- Saiju Pyarajan Ph.D., Director, Data and Computational Sciences
VA Boston Healthcare System, 150 S. Huntington Avenue, Boston, MA 02130
- Luis E. Selva, Ph.D., Executive Director, MVP Biorepositories
VA Boston Healthcare System, 150 S. Huntington Avenue, Boston, MA 02130
- Shahpoor (Alex) Shayan, M.S., Director, MVP Recruitment and Enrollment Informatics
VA Boston Healthcare System, 150 S. Huntington Avenue, Boston, MA 02130
- Stacey B. Whitbourne, Ph.D., Director, MVP Cohort Management
VA Boston Healthcare System, 150 S. Huntington Avenue, Boston, MA 02130
- MVP Coordinating Centers
    - MVP Coordinating Center, Boston - J. Michael Gaziano, M.D., M.P.H.
    VA Boston Healthcare System, 150 S. Huntington Avenue, Boston, MA 02130
    - MVP Coordinating Center, Palo Alto – Philip S. Tsao, Ph.D.
    VA Palo Alto Health Care System, 3801 Miranda Avenue, Palo Alto, CA 94304
    - MVP Information Center, Canandaigua – Brady Stephens, M.S.
    Canandaigua VA Medical Center, 400 Fort Hill Avenue, Canandaigua, NY 14424
    - Cooperative Studies Program Clinical Research Pharmacy Coordinating Center, Albuquerque – Todd Connor, Pharm.D.; Dean P. Argyres, B.S., M.S.
    New Mexico VA Health Care System, 1501 San Pedro Drive SE, Albuquerque, NM 87108

**MVP Publications and Presentations Committee**

- Co-Chair: Themistocles L. Assimes, M.D., Ph. D
VA Palo Alto Health Care System, 3801 Miranda Avenue, Palo Alto, CA 94304
- Co-Chair: Adriana Hung, M.D.; M.P.H
VA Tennessee Valley Healthcare System, 1310 24$^{th}$ Ave. South, Nashville, TN 37212
- Co-Chair: Henry Kranzler, M.D.
Philadelphia VA Medical Center, 3900 Woodland Avenue, Philadelphia, PA 19104

**MVP Local Site Investigators**

- Samuel Aguayo, M.D., Phoenix VA Health Care System
650 E. Indian School Road, Phoenix, AZ 85012
- Sunil Ahuja, M.D., South Texas Veterans Health Care System
7400 Merton Minter Boulevard, San Antonio, TX 78229
- Kathrina Alexander, M.D., Veterans Health Care System of the Ozarks
1100 North College Avenue, Fayetteville, AR 72703
- Xiao M. Androulakis, M.D., Columbia VA Health Care System
6439 Garners Ferry Road, Columbia, SC 29209
- Prakash Balasubramanian, M.D., William S. Middleton Memorial Veterans

Hospital
2500 Overlook Terrace, Madison, WI 53705
- Zuhair Ballas, M.D., Iowa City VA Health Care System
601 Highway 6 West, Iowa City, IA 52246-2208
- Elizabeth S. Bast, M.D., M.P.H., Miami VA Health Care System
1201 NW 16th Street, 11 GRC, Miami FL 33125
- Jean Beckham, Ph.D., Durham VA Medical Center
508 Fulton Street, Durham, NC 27705
- Sujata Bhushan, M.D., VA North Texas Health Care System
4500 S. Lancaster Road, Dallas, TX 75216
- Edward Boyko, M.D., VA Puget Sound Health Care System
1660 S. Columbian Way, Seattle, WA 98108-1597
- David Cohen, M.D., Portland VA Medical Center
3710 SW U.S. Veterans Hospital Road, Portland, OR 97239
- Louis Dellitalia, M.D., Birmingham VA Medical Center
700 S. 19th Street, Birmingham AL 35233
- Gerald Wayne Dryden, Jr., M.D., Ph.D., Louisville VA Medical Center
800 Zorn Avenue, Louisville, KY 40206
- L. Christine Faulk, M.D., Robert J. Dole VA Medical Center
5500 East Kellogg Drive, Wichita, KS 67218-1607
- Joseph Fayad, M.D., VA Southern Nevada Healthcare System
6900 North Pecos Road, North Las Vegas, NV 89086
- Daryl Fujii, Ph.D., VA Pacific Islands Health Care System
459 Patterson Rd, Honolulu, HI 96819
- Saib Gappy, M.D., John D. Dingell VA Medical Center
4646 John R Street, Detroit, MI 48201
- Frank Gesek, Ph.D., White River Junction VA Medical Center
163 Veterans Drive, White River Junction, VT 05009
- Michael Godschalk, M.D., Richmond VA Medical Center
1201 Broad Rock Blvd., Richmond, VA 23249
- Jennifer Greco, M.D., Sioux Falls VA Health Care System
2501 W 22nd Street, Sioux Falls, SD 57105
- Todd W. Gress, M.D., Ph.D., Hershel "Woody" Williams VA Medical Center
1540 Spring Valley Drive, Huntington, WV 25704
- Samir Gupta, M.D., M.S.C.S., VA San Diego Healthcare System
3350 La Jolla Village Drive, San Diego, CA 92161
- Salvador Gutierrez, M.D., Edward Hines, Jr. VA Medical Center
5000 South 5th Avenue, Hines, IL 60141
- Mark Hamner, M.D., Ralph H. Johnson VA Medical Center
109 Bee Street, Mental Health Research, Charleston, SC 29401
- John Harley, M.D., Ph.D., Cincinnati VA Medical Center
3200 Vine Street, Cincinnati, OH 45220
- Daniel J. Hogan, M.D., Bay Pines VA Healthcare System
10,000 Bay Pines Blvd Bay Pines, FL 33744
- Adriana Hung, M.D., M.P.H., VA Tennessee Valley Healthcare System
1310 24th Avenue, South Nashville, TN 37212
- Robin Hurley, M.D., W.G. (Bill) Hefner VA Medical Center
1601 Brenner Ave, Salisbury, NC 28144

- Pran Iruvanti, D.O., Ph.D., Hampton VA Medical Center
100 Emancipation Drive, Hampton, VA 23667
- Frank Jacono, M.D., VA Northeast Ohio Healthcare System
10701 East Boulevard, Cleveland, OH 44106
- Darshana Jhala, M.D., Philadelphia VA Medical Center
3900 Woodland Avenue, Philadelphia, PA 19104
- Seema Joshi, M.D., F.A.C.P., ABOIM; VA Eastern Kansas Health Care System
4101 S 4th Street Trafficway, Leavenworth, KS 66048
- Scott Kinlay, M.B.B.S., Ph.D., VA Boston Healthcare System
150 S. Huntington Avenue, Boston, MA 02130
- Michael Landry, Ph.D., Southeast Louisiana Veterans Health Care System
2400 Canal Street, New Orleans, LA 70119
- Peter Liang, M.D., M.P.H., VA New York Harbor Healthcare System
423 East 23rd Street, New York, NY 10010
- Suthat Liangpunsakul, M.D., M.P.H., Richard Roudebush VA Medical Center
1481 West 10th Street, Indianapolis, IN 46202
- Jack Lichy, M.D., Ph.D., Washington DC VA Medical Center
50 Irving St, Washington, D. C. 20422
- Tze Shien Lo, M.D., Fargo VA Health Care System
2101 N. Elm, Fargo, ND 58102
- C. Scott Mahan, M.D., Charles George VA Medical Center
1100 Tunnel Road, Asheville, NC 28805
- Ronnie Marrache, M.D., VA Maine Healthcare System Center, Augusta, ME 04330
- Stephen Mastorides, M.D., James A. Haley Veterans' Hospital
13000 Bruce B. Downs Blvd, Tampa, FL 33612
- Kristin Mattocks, Ph.D., M.P.H., Central Western Massachusetts Healthcare System
421 North Main Street, Leeds, MA 01053
- Paul Meyer, M.D., Ph.D., Southern Arizona VA Health Care System
3601 S 6th Avenue, Tucson, AZ 85723
- Jonathan Moorman, M.D., Ph.D., James H. Quillen VA Medical Center
Corner of Lamont & Veterans Way, Mountain Home, TN 37684
- Providencia Morales, R.N., Northern Arizona VA Health Care System
500 Highway 89 North, Prescott, AZ 86313
- Timothy Morgan, M.D., VA Long Beach Healthcare System
5901 East 7th Street Long Beach, CA 90822
- Maureen Murdoch, M.D., M.P.H., Minneapolis VA Health Care System
One Veterans Drive, Minneapolis, MN 55417
- Eknath Naik, M.D., Ph.D., West Palm Beach VA Medical Center,
7305 North Military Trail, West Palm Beach, FL 33410-6400
- James Norton, Ph.D., VA Health Care Upstate New York
113 Holland Avenue, Albany, NY 12208
- Olaoluwa Okusaga, M.D., Michael E. DeBakey VA Medical Center
2002 Holcombe Blvd, Houston, TX 77030
- Michael K. Ong, M.D., VA Greater Los Angeles Health Care System
11301 Wilshire Blvd, Los Angeles, CA 90073

- Kris Ann Oursler, M.D., Salem VA Medical Center
1970 Roanoke Blvd, Salem, VA 24153
- Ismene Petrakis, M.D., VA Connecticut Healthcare System
950 Campbell Avenue, West Haven, CT 06516
- Samuel Poon, M.D., Manchester VA Medical Center
718 Smyth Road, Manchester, NH 03104
- Amneet S. Rai, Pharm.D., VA Sierra Nevada Health Care System
975 Kirman Avenue, Reno, NV 89502
- Michael Rauchman, M.D., St. Louis VA Health Care System
915 North Grand Blvd, St. Louis, MO 63106
- Richard Servatius, Ph.D., Syracuse VA Medical Center
800 Irving Avenue, Syracuse, NY 13210
- Satish Sharma, M.D., Providence VA Medical Center
830 Chalkstone Avenue, Providence, RI 02908
- River Smith, Ph.D., Eastern Oklahoma VA Health Care System
1011 Honor Heights Drive, Muskogee, OK 74401
- Peruvemba Sriram, M.D., N. FL/S. GA Veterans Health System
1601 SW Archer Road, Gainesville, FL 32608
- Patrick Strollo, Jr., M.D., VA Pittsburgh Health Care System
University Drive, Pittsburgh, PA 15240
- Neeraj Tandon, M.D., Overton Brooks VA Medical Center
510 East Stoner Ave, Shreveport, LA 71101
- Philip Tsao, Ph.D., VA Palo Alto Health Care System
3801 Miranda Avenue, Palo Alto, CA 94304-1290
- Gerardo Villareal, M.D., New Mexico VA Health Care System
1501 San Pedro Drive, S.E. Albuquerque, NM 87108
- Jessica Walsh, M.D., VA Salt Lake City Health Care System
500 Foothill Drive, Salt Lake City, UT 84148
- John Wells, Ph.D., Edith Nourse Rogers Memorial Veterans Hospital
200 Springs Road, Bedford, MA 01730
- Jeffrey Whittle, M.D., M.P.H., Clement J. Zablocki VA Medical Center
5000 West National Avenue, Milwaukee, WI 53295
- Mary Whooley, M.D., San Francisco VA Health Care System
4150 Clement Street, San Francisco, CA 94121
- Peter Wilson, M.D., Atlanta VA Medical Center
1670 Clairmont Road, Decatur, GA 30033
- Junzhe Xu, M.D., VA Western New York Healthcare System
3495 Bailey Avenue, Buffalo, NY 14215-1199
- Shing Shing Yeh, Ph.D., M.D., Northport VA Medical Center
79 Middleville Road, Northport, NY 11768
- Andrew W. Yen, M.D., VA Northern California Health Care System
10535 Hospital Way, Mather, CA 95655
